# Antibiotic prescribing and resistance beyond hospital surveillance networks in India

**DOI:** 10.64898/2026.10.08.26365044

**Authors:** Jasmine Kaur, Vineet Madaan, Rajeev Sharma, Zoya Agarwal, Nachiket Gudi, Tikesh Bisen, Sachin Gupte, Ankur Mutreja, Nikhil Doegar, Harpreet Singh, Gaurav Agarwal, Tavpritesh Sethi

**Affiliations:** Center of Excellence in Healthcare, Indraprastha Institute of Information Technology Delhi, Okhla Industrial Estate Phase III, New Delhi, India, 110020; Department of Computational Biology, Indraprastha Institute of Information Technology Delhi, Okhla Industrial Estate Phase III, New Delhi, India, 110020; Tata 1mg Technologies Private Limited, Gurugram, Haryana, India; Centre for Health Research and Innovation (CHRI), a PATH affiliate, New Delhi, India; PATH, New Delhi, India; John Snow India Private Limited, New Delhi, India; Division of Development Research, Indian Council of Medical Research (ICMR), New Delhi, India

## Abstract

Hospital-based networks provide much of India’s antimicrobial resistance surveillance evidence, but community-facing data on both prescribing and resistance remain limited. We analysed 2.78 million prescription-validated antibiotic records and 155,764 microbiology samples from Tata 1mg, a digital healthcare platform, during 2023-2025. We assessed prescribing composition and resistance trends, and explored ecological prescribing-resistance associations using merged monthly series. Access antibiotics accounted for 28.7% of records, compared with 47.0% for Watch and 11.9% for Not Recommended antibiotics. Across 25 eligible states and union territories, Access shares ranged from 16.1% to 38.0%. Urine comprised 93.9% of samples. Among urinary Escherichia coli isolates, resistance was 4.9% to nitrofurantoin and amikacin, compared with 53.6% to ceftriaxone and 57.0% to ciprofloxacin. Exploratory prescribing-resistance associations varied by organism and antibiotic. These findings identify stewardship priorities within a digitally mediated care pathway and illustrate the complementary value of routinely collected pharmacy and laboratory data for surveillance beyond hospital-centred networks.

## Introduction

An estimated 1.14 million deaths worldwide were attributable to bacterial antimicrobial resistance (AMR) in 2021, with substantial variation by age and location^1^. AMR surveillance in India is supported by established hospital and laboratory networks, including the Indian Council of Medical Research Antimicrobial Resistance Research and Surveillance Network (ICMR AMRSN)^2^ and the National Centre for Disease Control’s National AMR Surveillance Network (NARS-Net)^3^. The ICMR AMRSN 2024 report documented 99,027 culture-positive isolates and described contributions from 21 regional centres^2^, while NARS-Net analysed 195,077 deduplicated priority bacterial isolates from 54 reporting laboratories^3^. These networks include hospital outpatients as well as admitted patients, but their findings reflect the populations accessing participating facilities and the clinical decisions to obtain cultures.

Alongside resistance surveillance, monitoring antibiotic prescribing can help identify stewardship priorities. The World Health Organization’s Access, Watch and Reserve (AWaRe) classification^4^ supports monitoring of antibiotic-use composition. The former WHO benchmark called for at least 60% of antibiotic consumption to comprise Access antibiotics^5^ and the 2024 United Nations political declaration on AMR established a target for Access antibiotics to account for at least 70% of global human antibiotic use by 2030^6^. In India, an analysis of private-sector antibiotic sales in 2019 estimated consumption of 5.07 billion defined daily doses, with Watch and Access antibiotics accounting for 54.9% and 27.0%, respectively^7^. Global analyses have also documented large geographic differences in antibiotic consumption and a disproportionate rise in Watch antibiotic use, particularly in low- and middle-income countries^8,9^. Prescribing composition and organism- and specimen-specific susceptibility describe different aspects of stewardship and examining them together can identify priorities for further review.

Digital healthcare platforms offering outpatient pharmacy and home-based diagnostic services provide an opportunity to examine these questions within a common care pathway. Prescription records can describe antibiotic prescribing, while cultures from home-collected urine specimens can characterise resistance among users of the diagnostic service. Their common platform infrastructure and observation period enable complementary characterisation of prescribing and resistance, although the two streams need not represent the same individuals or clinical episodes. Where geographic and temporal coverage overlaps, these data may also support locally focused stewardship review.

Here, we analysed routinely collected outpatient prescription records and diagnostic microbiology from specimens collected exclusively at home through Tata 1mg during 2023-2025. We characterised platform coverage, AWaRe prescribing composition, urinary resistance profiles and temporal trends, and contextualised resistance estimates against hospital-based surveillance reports. Exploratory analyses examined ecological lead-lag associations between prescribing and resistance using nationally pooled monthly series. Our objective was to evaluate how routinely collected prescribing and microbiology data can complement established AMR surveillance and identify stewardship priorities within the platform.

## Results

Between 2023 and 2025, the platform recorded 25.7 million prescribed medicine items and 155,764 home collected diagnostic microbiology samples. Antibiotics accounted for 2.78 million prescribed medicines (10.82%), with records spanning 35 states and union territories, 694 districts and 13,584 pincodes. State-specific prescription analyses included 25 states and union territories meeting the prespecified threshold of at least 1,000 antibiotic prescriptions. Diagnostic samples represented 23 states/UTs, 102 districts and 1,661 pincodes. The geographic distribution and monthly presence of both streams are shown in Figure 1, with dataset characteristics summarised in Table 1. In the microbiology data, urine cultures accounted for 93.9% of all diagnostic samples and 97% of culture-positive samples (Supplementary Table 1). Downstream resistance analyses were therefore done on urinary bacterial infections.

**Figure 1:**
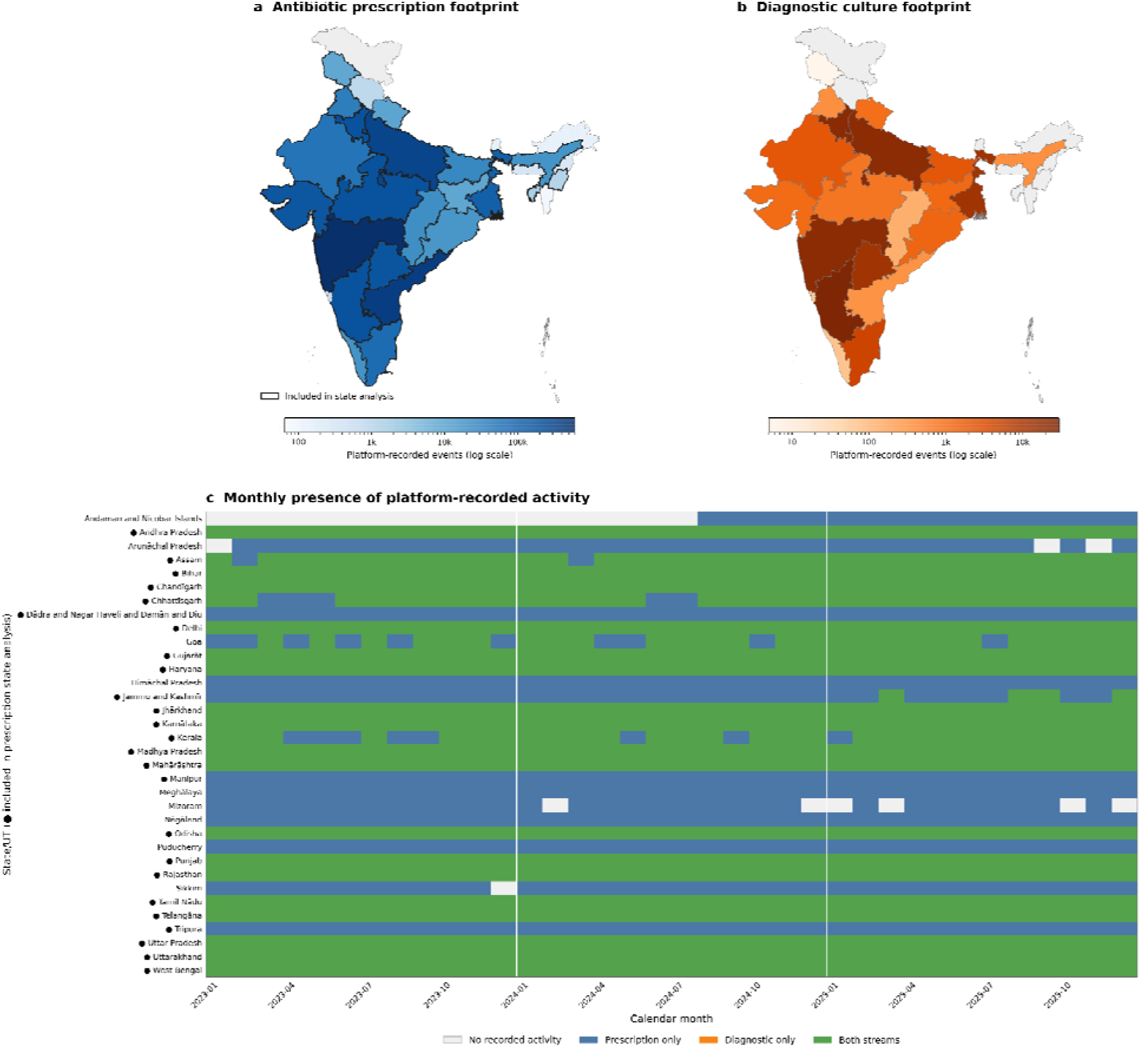
Geographic and temporal footprint of platform-recorded prescription and diagnostic activity across the datasets 2023-2025. (a) Antibiotic prescription activity across 35 source state/UT labels, corresponding to 34 current administrative boundary features; darker outlines identify the 25 states/UTs retained for state-specific analyses. (b) Diagnostic culture activity across 23 states/UTs. Grey areas indicate no recorded activity in the corresponding data stream. Fill intensity represents platform-recorded event counts on a logarithmic scale an should not be interpreted as population-level prescribing, resistance or platform penetration. (c) Monthly presence of prescription and diagnostic activity by state/UT.

**Table 1.** Scale, geographic coverage, and AWaRe composition of digital pharmacy datasets, India, 2023-2025.

| Domain | Measure | Prescription dataset | Diagnostic dataset |
|---|---|---|---|
| **Dataset size** | Total medicine recorded in all prescriptions | 25,735,715 | - |
|  | Antibiotic prescription records (>=1,000/state/UT) | 2,784,503; 2,778,086 | - |
|  | Antibiotics as a share of all prescribed medicines | 10.82% | - |
| **Geographic coverage** | States/UTs observed; state-specific eligible; below threshold | 35; 25; 10 (2,742 prescription medicines) | 23 |
|  | Districts represented, all prescriptions | 694 | 102 |
|  | Pincodes represented, all prescriptions | 13,584 | 1661 |
| **Temporal coverage** | Study period | 2023-2025 | 2023-2025 |
|  | Zero-activity state/UT-months among 35 valid geographies | 39/1,260; 3.10% | 57/828 state-months inactive |
|  | Active state/UT-month coverage among valid geographies | 1,221/1,260; 96.90% | 771/828; 93.12% |
| **State-month intensity** | Median prescriptions per active valid state/UT-month | 437 | - |
|  | IQR | 18-3,409 | - |
| **AWaRe composition** | Access | 798,643; 28.68% | - |
|  | Watch | 1,307,563; 46.96% | - |
|  | Reserve | 93,562; 3.36% | - |
|  | Not Recommended | 331,992; 11.92% | - |
|  | Unclassified | 252,743; 9.08% |  |
| **Gap from WHO Access target** | Gaps from former >=60%; 2030 >=70% Access targets | -31.32; -41.32 percentage points | - |
|  | Access-to-Watch ratio | 0.611 | - |
| **Dominant antibiotic classes** | Rank 1 | Beta-lactam/beta-lactamase-inhibitor: 589,954; 21.19% | - |
|  | Rank 2 | Macrolides: 569,675; 20.46% | - |
|  | Rank 3 | Third-generation cephalosporins: 432,635; 15.54% | - |
| **Diagnostic context** | Samples analysed | - | 155,764 |
|  | Culture-positive samples | - | 35,657; 22.89% |
|  | Urine among diagnostic samples | - | 146,273; 93.91% |
|  | Dominant culture-positive organism | - | Escherichia coli: 21,608; 60.60% |
| **Urine isolates** | *E. coli*-amoxicillin/clavulanic acid (Access) | - | 4,999/21,537; 23.21% |
|  | *E. coli*-nitrofurantoin (Access) | - | 1,060/21,539; 4.92% |
|  | *E. coli*-amikacin (Access) |  | 1,050/21,521; 4.88% |
|  | *E. coli*-ceftriaxone (Watch) |  | 10,499/19,594; 53.58% |
|  | *E. coli*-ciprofloxacin (Watch) |  | 12,275/21,542; 56.98% |
|  | *E. coli*-meropenem (Watch) |  | 307/10,635; 2.89% |
|  | *K. pneumoniae*-amoxicillin/clavulanic acid (Access) |  | 1,313/5,758; 22.80% |
|  | *K. pneumoniae*-nitrofurantoin (Access) |  | 1,382/5,754; 24.02% |
|  | *K. pneumoniae*-amikacin (Access) |  | 727/5,744; 12.66% |
|  | *K. pneumoniae*-ceftriaxone (Watch) |  | 1,853/5,751; 32.22% |
|  | *K. pneumoniae*-ciprofloxacin (Watch) |  | 1,911/5,755; 33.21% |
|  | *K. pneumoniae*-meropenem (Watch) |  | 313/3,827; 8.18% |

Values are n (%) unless otherwise stated. Medicines were classified by exact normalised whole-product matching to WHO AWaRe 2025; unmatched products were marked as Unclassified and used for the denominator. The former WHO Access benchmark was at least 60%, and the 2030 target is at least 70%. Resistance estimates are number of resistant isolates/tested isolates (%). Source data are from Supplementary Tables 1-3.

### AWaRe prescribing composition across states

Antibiotic prescriptions accounted for 10.82% of all medicine prescriptions, with monthly estimates ranging from 8.89% to 13.85% (Supplementary Figure 1). Overall, Watch antibiotics accounted for 1,307,563 prescriptions (46.96%), followed by Access 798,643 prescriptions (28.68%), Reserve for 93,562 (3.36%), Not Recommended combinations for 331,992 (11.92%), and Unclassified products for 252,743 (9.08%) (Table 1; Supplementary Table 3; Supplementary Figures 2-3). Access shares varied widely from approximately 16.1% in Chandigarh to 38.0% in Tamil Nadu. All eligible states/UTs had recorded Access shares below the former 60% and the 2030 70% reference benchmarks^5,6^ (Figure 2).

**Figure 2:**
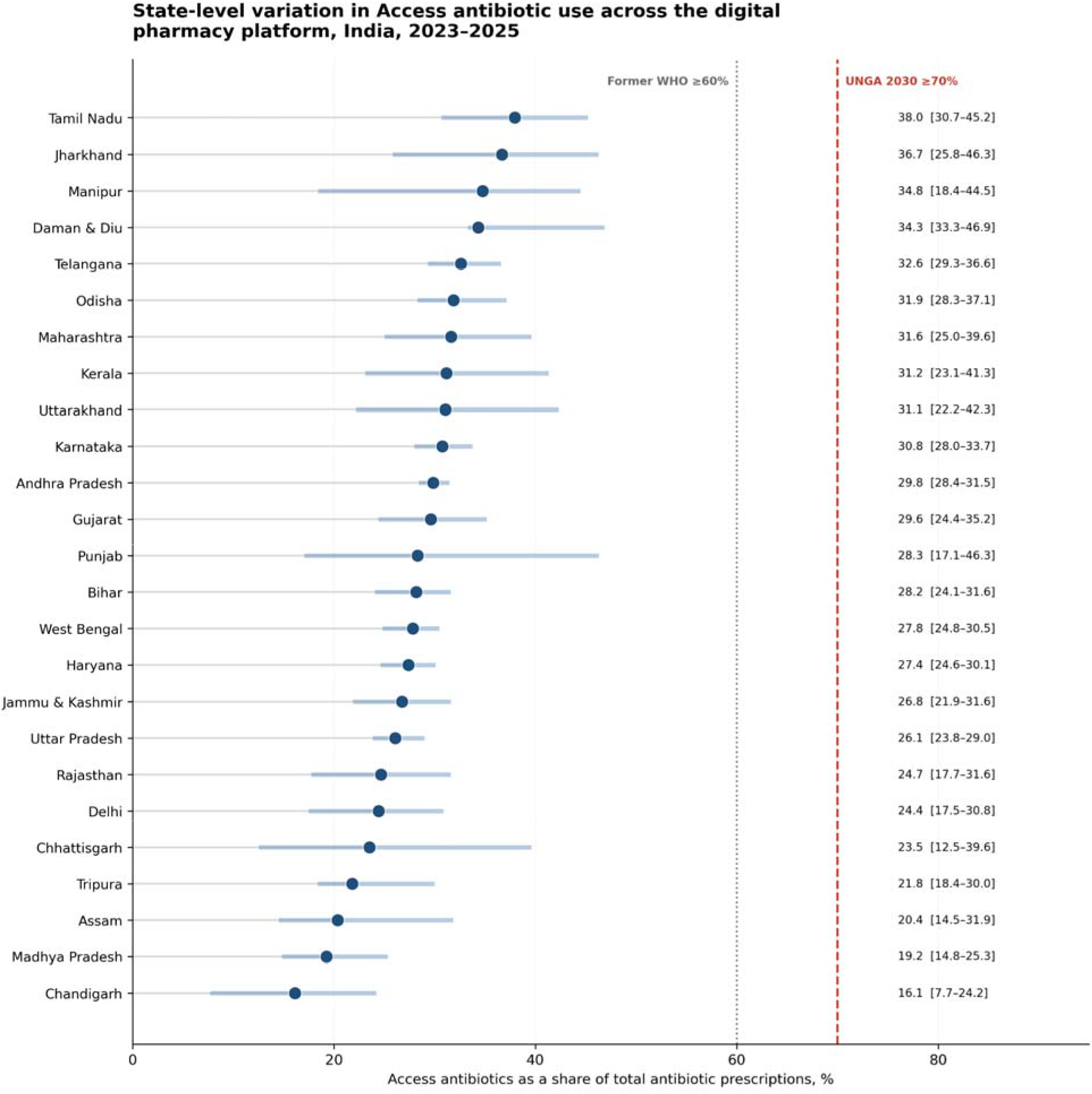
State-level variation in Access antibiotic use across the digital pharmacy platform, India, 2023-2025. State/UT-level proportions of Access antibiotic prescriptions among the 25 states/UTs meeting the prespecifie minimum of 1,000 antibiotic prescriptions. Products unmatched to a WHO AWaRe 2025 group remained in the denominator as Unclassified. Reference lines show the former 60% benchmark and the 2030 70% target. No eligible state/UT reached either threshold.

### Resistance varied substantially among antibiotics within AWaRe groups

Microbiology data contextualised this imbalance. Among 34,584 culture-positive urine samples, *E. coli* accounted for 21,549 isolates (62.31%), followed by *K. pneumoniae* with 5,759 (16.65%) and *P. aeruginosa* with 1,629 (4.71%) (Supplementary Table 1).

Among urinary *E. coli* isolates, resistance was 4.92% to nitrofurantoin and 4.88% to amikacin, compared with 53.58% to ceftriaxone and 56.98% to ciprofloxacin. Resistance also varied among Access antibiotics, reaching 23.21% for amoxicillin/clavulanic acid, 40.68% for trimethoprim/sulfamethoxazole and 82.80% for ampicillin (Table 1; Supplementary Table 2). Urinary K. pneumoniae showed a different resistance profile: resistance was 24.02% to nitrofurantoin, 12.66% to amikacin, 32.22% to ceftriaxone and 33.21% to ciprofloxacin. The number of isolates tested varied by antibiotic; resistant and tested counts are provided in Supplementary Table 2.

A comparison of *E. coli* urinary isolates in TATA 1mg data and the published outpatient estimates in NCDC NARS-Net 2024 report, revealed lower resistance estimates for nitrofurantoin, amikacin, ceftriaxone and ciprofloxacin (Figure 3). However, estimates were similar for ampicillin (83.4% versus 84%) and cefepime (42.0% versus 43%). Since the ICMR AMRSN report did not provide a corresponding urinary outpatient stratum direct comparison could not be done. In a separate comparison, resistance in Tata 1mg urinary *K. pneumoniae* isolates was lower than that reported for NCDC urinary *Klebsiella* spp. across all care settings, including ceftriaxone (27.0% versus 68%) and ciprofloxacin (27.1% versus 61%).

**Figure 3:**
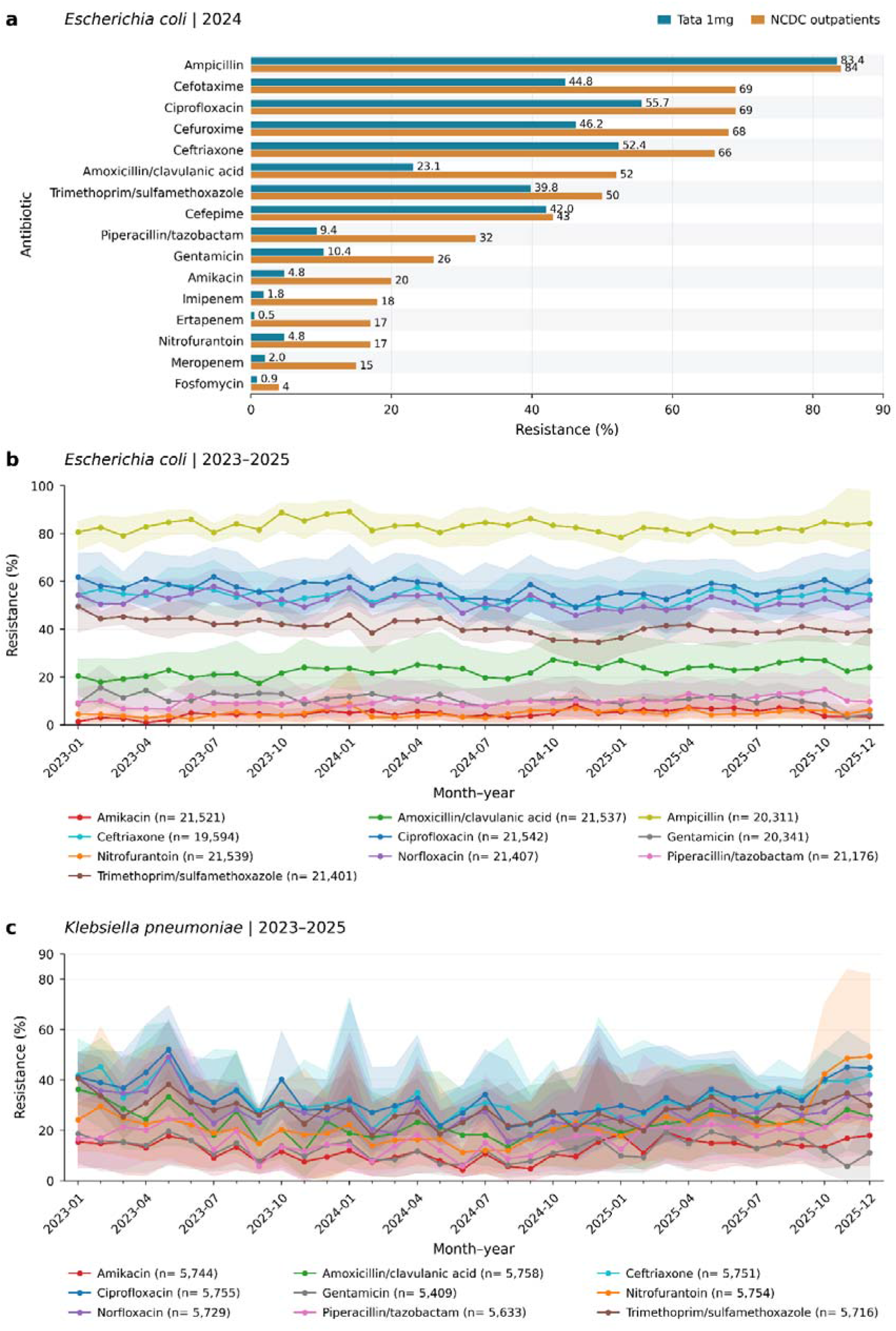
Urinary antimicrobial resistance in Tata 1mg data and comparison with hospital-outpatient surveillance. a. Resistance among urinary *Escherichia coli* isolates in Tata 1mg data and the National Centre for Disease Control (NCDC) hospital-outpatient surveillance data in 2024. Values beside bars indicate resistance percentages. Comparisons are descriptive and unadjusted for differences in patient populations, sampling and testing practices. (b) Monthly resistance among top 10 antibiotics for urinary *E. coli* and (c) top nine antibiotics for *Klebsiella pneumoniae* isolates from January 2023 to December 2025. Points and lines represent observed monthly resistance proportions and shaded bands indicate 95% percentile bootstrap confidence intervals from 1,000 resamples of states within each month-antibiotic group.

In descriptive pincode level analysis, prescribing share and urinary resistance were compared between 2023 and 2025 for 11,493 pincode-organism-antibiotic combinations across 648 pincodes. Both measures increased in 921 comparisons across 374 pincodes, involving 19 antibiotics (622 comparisons for *E. coli* and 299 for *K. pneumoniae*). Both measures decreased in 850 comparisons, changed in opposite directions in 1,758 comparisons. Among 9,795 comparisons with both measures available in all three years, 78 comparisons across 63 pincodes showed successive annual increases during 2023-2025 (Supplementary Table 4).

### Temporal trends in urinary antimicrobial resistance

Among urinary *E. coli*, statistically significant increase was observed for amoxicillin/clavulanic acid 0.172 percentage points per month (BH-adjusted p = 0.0010), piperacillin/tazobactam 0.131 percentage points per month (BH-adjusted p = 0.0080), amikacin 0.097 percentage points per month (BH-adjusted p = 2.3 × 10^−4^) and nitrofurantoin 0.061 percentage points per month (BH-adjusted p = 0.0075). Resistance declined significantly for cefepime, cefotaxime, ceftazidime, cefuroxime, trimethoprim-sulfamethoxazole, ofloxacin, ticarcillin, norfloxacin and gentamicin (Supplementary Table 5; Figure 3b).

Among urinary *K. pneumoniae*, statistically significant increase was observed for nitrofurantoin resistance at 0.398 percentage points per month (BH-adjusted p = 1.0 × 10^−6^). Resistance to tetracycline, cefepime and ofloxacin declined significantly (Supplementary Table 6; Figure 3c).

Ertapenem resistance showed significant positive model-based trends in both urinary *E. coli* and *K. pneumoniae*, with higher observed resistance particularly evident from May 2025 onward (Supplementary Tables 5-6).

No antibiotic showed a significant longitudinal trend for urinary *Pseudomonas aeruginosa* after multiple-testing correction (Supplementary Table 7; Supplementary Figure 4)

### Temporal trends in antibiotic prescribing

Although the overall AWaRe composition remained broadly stable, individual antibiotics showed changes in their share of recorded antibiotic prescriptions (Figure 4). Model-based estimates reveal that within Access antibiotics, clindamycin increased by 0.063 percentage points per month (BH-adjusted p = 2.4 × 10^−35^), tetracycline by 0.002 (BH-adjusted p = 5.6 × 10^−4^) and sultamicillin by 0.002 (BH-adjusted p = 0.0026). Amikacin and benzylpenicillin declined by

**Figure 4.**
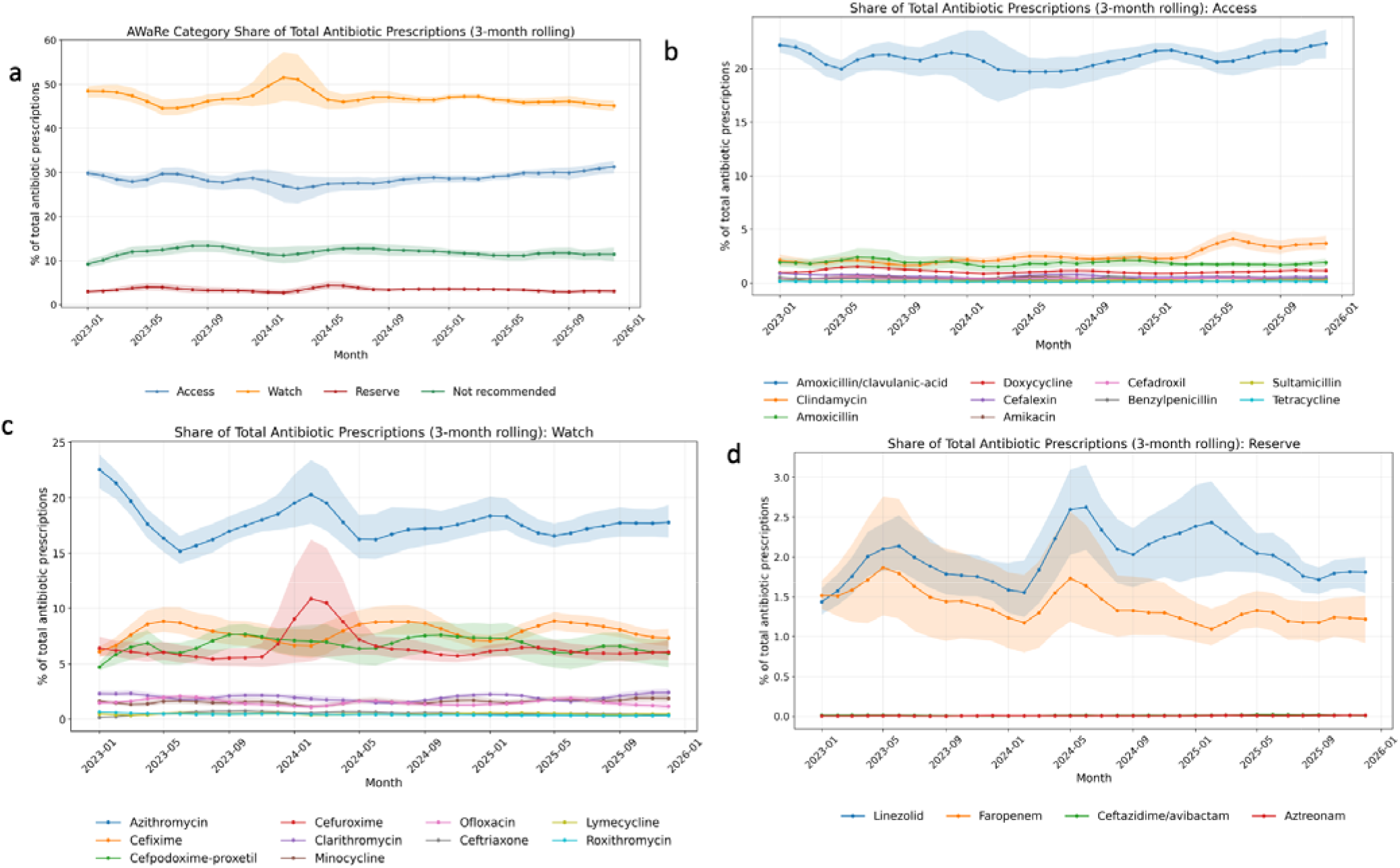
Temporal trends in antibiotic prescriptions by AWaRe category and drug, 2023-2025. (a) Trends of total antibiotic prescriptions classified as Access, Watch, or Reserve. (b-d) Antibiotic-level trends of total prescriptions within Access, Watch, and Reserve categories. Watch antibiotics remained the dominant prescription category throughout most of the study period, while Access antibiotics stayed consistently below the WHO 60% benchmark and Reserve antibiotics contributed a small share of total prescriptions.

0.004 percentage points per month (BH-adjusted p = 1.8 × 10^−6^ and 0.035). Among Watch antibiotics, minocycline and lymecycline increased by 0.025 (BH-adjusted p = 2.5 × 10^−13^) and 0.004 (BH-adjusted p =0.005) percentage points per month. Azithromycin, cefuroxime, clarithromycin and roxithromycin declined by 0.073, 0.031, 0.008 and 0.004 percentage points per month, respectively (all BH-adjusted p < 0.05). Among Not Recommended combinations, cefpodoxime proxetil/clavulanic acid increased by 0.023 percentage points per month (BH-adjusted p = 1.1 × 10^−4^), whereas cefixime/ofloxacin, cefixime/clavulanic acid, cefpodoxime proxetil/ofloxacin and amoxicillin/dicloxacillin declined (all BH-adjusted p < 0.05). Reserve prescribing remained low, although ceftazidime/avibactam and aztreonam increased by 0.0002 and 0.0002 percentage points per month, respectively (BH-adjusted p = 0.019 and 4.0 × 10^−4^).

In the sensitivity analysis using all medicine prescriptions as the denominator (Supplementary Table 8-9), all 18 significant trends retained their direction, and 16 remained significant after correction. The increases in lymecycline and cefpodoxime proxetil/clavulanic acid were no longer significant (BH-adjusted p = 0.497 and 0.241, respectively), whereas the increase in minocycline remained significant (BH-adjusted p = 9.5 × 10^−6^). Ten additional antibiotics or combinations showed significant declines only under the all-medicine denominator: amoxicillin/clavulanic acid, doxycycline, cefalexin, cefadroxil, cefoperazone/sulbactam, linezolid, faropenem, cefixime, ofloxacin and ceftriaxone.

### Lead-lag patterns in antibiotic resistance

In urinary *E. coli*, cross-correlation analysis identified repeated lagged associations in which Watch-class resistance preceded Access-class resistance by approximately 3-6 months.

Resistance to ceftazidime and ceftriaxone preceded resistance to trimethoprim-sulfamethoxazole by approximately 4 months, while ceftriaxone resistance also preceded tetracycline and ampicillin resistance by approximately 3-6 months (Supplementary Figure 5). These examples suggest temporal coupling between selected Access-class resistance signals and preceding Watch-class resistance signals, but they should be interpreted as exploratory surveillance markers and need prospective validation.

### Lead-lag patterns in antibiotic resistance and prescription antibiotics

Prewhitened cross-correlation analysis was performed for 18 antibiotic-stream pairs per organism, separately comparing normalised prescription counts with resistance rates.

For urinary *Klebsiella pneumoniae*, we observed an inverse association between resistance and subsequent antibiotic use. Several Watch antibiotics showed negative lagged correlations, where higher resistance was followed by lower prescription of the same antibiotic. This pattern was observed for ceftazidime, where resistance preceded lower prescribing at 4, 8 and 9 months and levofloxacin where resistance preceded lower prescribing at 2 months (Supplementary Figure 6). These findings are compatible with a shift away from antibiotics with rising resistance in *K. pneumoniae* consistent with therapeutic substitution or prescriber adaptation rather than a simple use-driven selection pattern. For aztreonam, prescription-leading-resistance signals were observed at 1 month and amoxicillin/clavulanic acid at 3 months (Supplementary Figure 7). Given the exploratory nature of cross-correlation analysis, these signals were interpreted as temporal surveillance markers rather than causal effects.

For urinary *Escherichia coli*, temporal patterns were more heterogeneous and did not suggest a single dominant organism-level pattern. A limited number of use-leading-resistance signals were observed: cefotaxime prescriptions preceded cefotaxime resistance by approximately 3 months (Supplementary Figure 8). Overall, unlike *K. pneumoniae*, the *E. coli* signals were more antibiotic-specific and less directionally coherent, suggesting heterogeneous temporal coupling between community antibiotic use and resistance.

## Discussion

This study analyses routinely collected digitised prescriptions and diagnostic microbiology data from a large digital healthcare platform in India, providing a community-facing view of antibiotic use and resistance that is currently missing in the conventional hospital-based AMR surveillance. To our knowledge, this is the first study to examine both antibiotic prescribing and microbiological resistance using routinely generated data from a digital healthcare platform in India. The study identifies a low Access prescribing share across eligible platform geographies, substantial variation in urinary resistance profiles, and changes over time that were not apparent from published reports alone. The analysis of prescription and resistance from the same platform also enabled exploratory assessment of temporal associations and local co-occurrence of prescribing and resistance changes.

Across all the eligible geographies, access antibiotics accounted for 28.7% of recorded antibiotics, compared with 47.0% for Watch antibiotics and 11.9% for Not Recommended combinations. The low Access share was geographically widespread and every state and union territory contributing at least 1,000 antibiotic records remained below the former 60% reference benchmark. This pattern is broadly consistent with the previously published results from India. Koya and colleagues reported 5.07 billion defined daily doses of systemic antibiotics consumed in India in 2019, with Watch antibiotics accounting for 54.9% and Access antibiotics for only 27.0% of total consumption^7^. Farooqui and colleagues similarly showed increasing community-level antibiotic consumption in India, along with growing use of newer and broader-spectrum classes^10^. Internationally, Klein and colleagues found that per-capita Watch consumption increased by 90.9% between 2000 and 2015, compared with 26.2% for Access antibiotics, with a larger increase in Watch use in low- and middle-income countries^9^. Our record-based estimates are not directly comparable with defined daily doses, but both studies identify a predominance of Watch antibiotics. Browne and colleagues estimated that antibiotic consumption rates in South Asia increased by 116% between 2000 and 2018, alongside substantial national and subnational variation in antibiotic use^8^. These findings identify an important stewardship opportunity within the platform, although they do not establish whether individual prescriptions were clinically appropriate. The WHO Access targets are population-level benchmarks, whereas the WHO AWaRe antibiotic book provides guidance by infection syndrome, including antibiotic choice, dose, route and duration^11^. Increasing Access use therefore requires consideration of diagnosis, susceptibility and patient characteristics.

The resistance findings also show why stewardship cannot be reduced to increasing the Access share alone. Among urinary *E. coli*, resistance varied substantially within the Access group, from relatively low levels for nitrofurantoin and amikacin to substantially higher levels for trimethoprim/sulfamethoxazole and ampicillin. *K. pneumoniae* showed a different profile, including higher nitrofurantoin resistance. Antibiotic selection therefore requires consideration of the organism, infection site, disease severity, route of administration and patient characteristics alongside AWaRe classification and local susceptibility evidence^12–13^.

The urinary resistance profiles observed through Tata 1mg provide context alongside established hospital-based surveillance. The comparison restricted to urinary *E. coli* for OPD cases in NCDC 2024 report^3^ illustrates the complementary information provided by the platform. Resistance proportions were lower than NCDC hospital-outpatient estimates for several antibiotics, including nitrofurantoin, amikacin, ceftriaxone and ciprofloxacin, whereas estimates for ampicillin and cefepime were similar. NCDC provided a corresponding urinary outpatient stratum and the relevant setting-stratified tables in the ICMR report excluded urine. Restricting the comparison by year, organism, specimen and resistance endpoint improved its interpretability, but patient characteristics, previous treatment, geographic composition, culture selection and laboratory practices could not be harmonised. The differences therefore cannot be attributed to the care pathway itself and shows that surveillance through another care pathway can yield a different observed resistance profile, even when the broad organism and specimen definitions are aligned.

Longitudinal findings add an important dimension to these comparisons. Despite relatively low pooled resistance in *E. coli*, nitrofurantoin resistance increased over time in both *E. coli* and *K. pneumoniae*. Amikacin resistance also increased in *E. coli*, alongside increases for amoxicillin/clavulanic acid and piperacillin/tazobactam. Several other antibiotics showed declining resistance, indicating that changes were specific to organism-antibiotic combinations rather than uniformly worsening. Our earlier multicentre analysis of urinary infections from the ICMR AMRSN network (2017-2021) also identified increasing nitrofurantoin resistance in *E. coli*, although the populations and time period differed^14^. Catalán and colleagues similarly identified heterogeneous temporal patterns across pathogen-antibiotic pairs in the global ATLAS database, including methodological artefacts that could alter apparent trends^15^. These findings support considering resistance levels and trajectories together within a geography as a relatively low pooled proportion can coexist with an increase that warrants continued monitoring. A descriptive pincode level analysis provided a more local view of the two data streams.

Our exploratory lead-lag analyses revealed that for urinary *E. coli* resistance to Watch-class antibiotics preceded resistance to Access-class antibiotics by approximately 3-6 months for several combinations. Ceftazidime and ceftriaxone resistance preceded trimethoprim-sulfamethoxazole resistance by around 4 months, while ceftriaxone resistance preceded tetracycline and ampicillin resistance by approximately 3-6 months. These findings are conceptually aligned with our earlier indicator-antibiotic framework in which some antibiotics showed increasing patterns earlier relative to critically important antibiotics and could be used as potential early warning signals^14,16^. A systematic review of hospitalised adults in Europe found that most reported associations between antibiotic use and resistance occurred within 0-6 months^17^. In the present platform dataset, the lead-lag structure suggests that selected Watch-resistance trends may warrant prospective evaluation as candidate leading indicators, using a prespecified lag and an independent dataset. However, these associations are not causal and several factors like co-selection, plasmid linkage, clonal expansion, diagnostic selection, seasonality, changing case mix, and antibiotic exposure outside the platform may all contribute. Several limitations affect interpretation of these findings. The data represent platform-mediated healthcare interactions and were not sampled to represent the Indian population. Findings should therefore be interpreted within the online digital healthcare platform. Since urine predominated the diagnostic samples, the downstream resistance analyses was done on urinary isolates. Although the platform contains home-collected samples, collection location does not establish where infection was acquired or exclude previous hospital exposure. Therefore, the resistance estimates should therefore not be interpreted as estimates of community-acquired resistance. Patients selected for culture may differ from other platform users in previous antibiotic treatment, recurrence, disease complexity or treatment failure. The aggregated diagnostic records lacked prior-antibiotic exposure, clinical indications and repeat-testing identifiers, so we could not quantify these differences or assess the contribution of repeated samples from the same patients. In an independent survey of users of the same platform, 46.7% of respondents in the doctor-recommended antibiotic subgroup reported that a doctor had recommended a diagnostic test before prescribing^18^. This finding provides context for diagnostic use within the platform population, although recommendations for any diagnostic test do not establish the frequency of culture testing or the reasons patients were selected for culture. Resistance proportions should therefore be interpreted as describing the tested isolates, rather than the broader platform outpatient population. The independent survey also found that 58.0% of respondents reported that their last antibiotic use was doctor-recommended, while 42.1% reported always completing the full antibiotic course^18^. These findings highlight variation in how antibiotics are obtained and taken, which prescription records alone cannot capture. However, these self-reported practices cannot be used to quantify antibiotic consumption or adherence in the analysed prescription cohort. Recorded prescriptions should therefore be interpreted as a measure of antibiotic activity through the platform, rather than a direct measure of individual antibiotic exposure.

Since aggregated data was analysed, the individual-level prescriptions and diagnostic results could not be linked. Their common platform origin therefore does not establish that patients contributing diagnostic samples were exposed to the antibiotics recorded in the prescription dataset. Prescription–resistance associations should consequently be interpreted as ecological associations between recorded prescribing and resistance, without attributing a subsequent resistant isolate to a particular prescription or purchase.

The time-series analyses also have limitations. Cross-correlation identifies temporal ordering but cannot distinguish direct selection from co-selection, seasonal disease patterns, local outbreaks, shifting diagnostic intensity, or changes in platform coverage. We used prewhitening to reduce the influence of serial correlation, but lagged associations may still reflect co-selection, seasonal disease patterns, local outbreaks, changes in diagnostic testing or shifts in platform coverage. These signals should therefore be considered exploratory and require independent validation before use as early warning indicators.

Despite these limitations, the study demonstrates that routine digital health data from an online pharmacy platform can be repurposed into a high-frequency AMR intelligence layer. It will not replace nationally representative consumption audits or formal AMR surveillance networks but may add a complementary, operational view of a community-linked care pathway in which stewardship interventions may be deployable through prescription review, pharmacy workflows, diagnostic prompts, state-specific feedback, and digital nudges. Future direction will involve determining whether these signals remain stable within consistently observed populations and whether prospectively generated alerts provide information early enough to influence stewardship decisions. In the absence of nation-wide community data, digital healthcare platforms can provide a useful operational layer alongside established national AMR surveillance in India.

## Methods

### Study Design

We conducted a retrospective observational time-series study using routinely collected digitised prescription and diagnostic microbiology data from Tata 1mg, a large digital healthcare platform in India. Prescription records contained aggregated by antibiotic, state and month, while diagnostic data contained aggregated by organism, specimen type, antibiotic, state, pincode and month. The two streams are from a shared digitally mediated outpatient care pathway (TATA 1mg) but were analysed as aggregated records without individual patient level linkage. Records generated between 1 January 2023 and 31 December 2025 were included for analysis. Antibiotic names in both the datasets were standardised and mapped to WHO AWaRe 2025 classification^4^. The prescription cohort included all prescription-validated antibiotics recorded during the study period with a valid prescription date and a positive prescription count. Products that did not represent antibacterial agents were excluded from the downstream analysis. Products without a corresponding entry in the WHO AWaRe 2025 lists were retained as Unclassified.

To quantify antibiotic activity relative to overall platform use, monthly prescription antibiotic counts were expressed as a proportion of all prescription medicines recorded during the corresponding month. In contrast, analyses of AWaRe composition and individual antibiotic trends used total antibiotic prescriptions within the relevant month or state-month as the denominator. These proportional measures reduced the influence of secular changes in platform activity and enabled comparison of prescribing patterns over time.

All eligible prescription antibiotics were included in national analyses. State-level analyses were restricted to states and union territories with a valid location and at least 1,000 prescription antibiotics during the study period.

The diagnostic dataset included diagnostic specimens were collected exclusively from patients’ homes and processed in Tata 1mg laboratories during the study period. Resistance analyses were restricted to culture-positive bacterial isolates and downstream analysis were restricted to culture-positive urine samples. Since aggregated data were analysed no attempt was made to re-identify any patient.

### Data sources

This study comprehensively analysed a novel dataset from Tata 1mg, a large online pharmacy and digital health platform that has operated in India since 2015. Routinely generated aggregated data for antibiotic prescriptions and diagnostic microbiology were analysed for the period 2023 to 2025.

#### Prescription dataset

The antibiotic prescription data were derived from orders placed through the Tata 1mg platform that required a valid prescription. Under the platform workflow, users upload prescriptions to purchase medicines, which are then digitised, reviewed, and confirmed by an internal team of experts. The medicine names and corresponding antibiotic names from these prescriptions were at a pincode and day-level for the analysis.

#### Diagnostic dataset

The diagnostic data pertaining to the culture results were obtained from Tata 1mg laboratories located across 18 Indian cities: Delhi, Gurgaon, Lucknow, Dehradun, Jaipur, Mumbai, Pune, Ahmedabad, Bangalore, Chennai, Hyderabad, Bhubaneswar, Angul, Ranchi, Patna, Kolkata, Guwahati, and Visakhapatnam. These laboratories process clinical specimens, including urine and blood samples, and generate microbiological reports containing organism identification and antimicrobial susceptibility testing results. The diagnostic dataset used in this study included aggregated data for isolated organisms and the antibiotic susceptibility profiles. Microbiological testing was performed using the VITEK 2 Compact system, based on colourimetric and turbidimetric principles for rapid organism identification and susceptibility testing. Laboratory workflows were organised by pathogen group using GPID, GNID, and YST identification cards, together with organism-appropriate AST cards, including AST-P628 for Gram-positive organisms and AST-N405, AST-N477, and AST-N406 for Gram-negative organisms. The interpretation of antimicrobial susceptibility testing results was based on the Clinical and Laboratory Standards Institute guidelines in use at the time of reporting. Internal quality control procedures were routinely maintained using American Type Culture Collection reference strains. Weekly bacterial quality control included *Enterococcus faecalis* ATCC 29212, *Staphylococcus aureus* ATCC 29213, *Escherichia coli* ATCC 25922, and *Pseudomonas aeruginosa* ATCC 27853. Fungal quality control for *Candida albicans* and *Candida parapsilosis* was conducted monthly. External quality assurance was supported through participation in the IAMM programme, coordinated by Sir Ganga Ram Hospital, every four months, and in CAP proficiency testing every three months.

These data sources enabled integration and analysis of antibiotic use and antimicrobial resistance patterns within a large digitally mediated, community-linked healthcare setting in India.

### Data Pre-processing

Diagnostic specimen labels were harmonised into urine, sputum and blood categories. Organism names were cleaned by removing specified report annotations, standardising spelling and taxonomic abbreviations, and applying explicit mappings, longest-prefix matching and, where necessary, approximate matching against a reference vocabulary using a similarity threshold of 0.75. Entries consisting of colony-count information rather than organism names were treated as missing, and recognised negative-culture descriptions were standardised as No growth. Laboratory antibiotic names were cleaned to remove embedded codes, susceptibility descriptors and extraneous characters, with explicit rules for spelling variants and combination agents. Antibiotics in both datasets were classified using the WHO AWaRe 2025 classification^4^. For prescription data, antibiotic names were standardised for case, spacing, component separators and recognised generic-name variants. Fixed-dose combinations were classified only when the complete combination matched a WHO entry. Medicine names were first matched to the Not Recommended list and subsequently to Access, Watch and Reserve groups. Medicines that could not be matched were retained as Unclassified.

For the diagnostic dataset, antibiotic names were standardised using manually reviewed spelling corrections.

All prescription medicines were included in national analyses. State-specific prescription analyses were restricted to the 25 states and union territories contributing at least 1,000 prescription antibiotics.

### Descriptive analysis

Descriptive analyses were performed separately for the diagnostic and prescription datasets. For each dataset, the geographic coverage was summarised by states and districts represented, and temporal coverage by study months and observed state-month activity. In the diagnostic dataset, we described culture outcomes, specimen types, organism distributions, and state-level contributions using counts and percentages. Resistance was summarised for major organism-antibiotic combinations as the proportion of resistant isolates among tested isolates, as in previous studies^14,16,19^. Monthly resistance proportions were displayed without smoothing, with 95% percentile bootstrap intervals derived from 1,000 resamples of states within each organism-antibiotic-month combination. For each organism, figures included the ten antibiotics with the largest number of interpretable susceptibility results.

The prescription dataset was summarised by total antibiotics, geographic and temporal coverage, top contributing states, and proportional distributions across AWaRe categories and antibiotic classes. Monthly prescription shares were displayed as centred three-month moving averages, with 95% percentile bootstrap intervals derived from 1,000 resamples of states within each month and the same smoothing applied to each bootstrap series.

### Comparison with published surveillance network reports

To contextualise platform-derived resistance estimates, we compared resistance among urinary *Escherichia coli* isolates collected through Tata 1mg during 2024 with the corresponding hospital-outpatient estimates reported in the National Centre for Disease Control’s NARS-Net 2024 annual report^3^. An equivalent comparison with ICMR AMRSN was not possible because its 2024 report did not provide outpatient-specific susceptibility estimates for urinary *E. coli*^2^. Comparisons were restricted to antibiotics reported in both datasets. These comparisons were descriptive and differences in patient characteristics, geographic coverage, culture selection, isolate deduplication and laboratory testing practices were not harmonised or adjusted for.

### Longitudinal trends for antibiotic prescriptions and resistance

Trends in antibiotic prescriptions and resistance were estimated using state-level generalised linear mixed models fitted to monthly count data. Models were fitted using the glmmTMB package^20^.

Prescription trends were analysed using beta-binomial generalised linear mixed models with a logit link, with the proportion of prescriptions attributable to each antibiotic as the dependent variable, time in months as a fixed-effect predictor, and state-level random intercepts and slopes. Separate models were fitted for up to ten most frequently prescribed products within each of the Access, Watch, Reserve and Not Recommended categories. State-months with antibiotic activity but no recorded prescriptions for the selected product were assigned a zero numerator. States with antibiotic activity in at least 75% of study months were included. If the initial prescription model failed to converge, a random-intercept-only beta-binomial model was attempted, followed by binomial models with random intercepts and slopes and then random intercepts only. Temporal effects were expressed as average marginal changes in antibiotic share, reported in percentage points per month, and p-values were adjusted within each AWaRe category using the Benjamini-Hochberg method^21^. Sensitivity analyses using all medicine prescriptions as the denominator was performed.

Resistance trends were analysed for urine isolates, specifically for each organism-antibiotic combination using beta-binomial generalised linear mixed models with a logit link, with the resistance percentage as the dependent variable, time in months as a fixed-effect predictor, and state-level random intercepts and slopes. For each organism-antibiotic combination, states were included if they had observations in at least 75% of the calendar months between the first and last observations for that combination and represented at least two pincodes. If the initial model could not be fitted successfully, a binomial model with the same random-effects structure was attempted, followed by random-intercept-only beta-binomial and binomial models. Temporal effects were summarised as average conditional marginal changes in antibiotic prescribing share or resistance, expressed in percentage points per month. Fitted probabilities incorporated state-level random intercepts and, where present, random slopes. For each observed state-month, the time derivative of the fitted probability was calculated using the common time coefficient plus the state-specific random slope and the latter was set to zero for random-intercept-only models. Derivatives were converted to percentage points and averaged with equal weight across the observed state-months included in each model. p-values were adjusted using the Benjamini-Hochberg method within each AWaRe category, separately for each prescription denominator, and within each organism for resistance.

### Integrating antibiotic prescriptions and resistance datasets

Antibiotic prescription and diagnostics datasets obtained from the Tata 1mg platform were merged to generate a unified monthly analytical dataset for downstream temporal analyses. The diagnostics dataset was filtered to include only culture-positive urine samples. For each organism-antibiotic combination, the resistance percentage was calculated as the proportion of resistant isolates among all isolates tested. The prescription dataset was independently processed and aggregated at the month and antibiotic-level to obtain normalised monthly prescription volume. Following aggregation, the prescription and diagnostic datasets were aligned using a common temporal and antibiotic framework. Specifically, pincode-wise resistance estimates for each organism-antibiotic combination in a given month were linked with the corresponding monthly prescription count for the same antibiotic and month. Since the microbiology data were retained at the organism level, whereas prescription data were available at the antibiotic level, the merged dataset preserved organism-specific resistance observations and antibiotic-specific prescription for the corresponding month and geography. Thus, the resulting linkage was temporal and platform-level rather than patient-level, and no individual record linkage between prescriptions and diagnostic results was performed.

### Lead-lag analyses of prescription and resistance

Lead-lag relationships were assessed using merged monthly time series of antibiotic prescriptions and organism-specific resistance. Analyses were performed separately for each organism-antibiotic pair. Antibiotic prescriptions were expressed as normalised monthly shares, and resistance was expressed as the monthly proportion of resistant isolates among those tested for that antibiotic. A complete calendar-month grid was retained for each series pair. Missing months remained missing and were not interpolated, ensuring that reported lags represented true calendar intervals. At least 24 paired months were required. The prescription series was prewhitened using an automatically selected ARIMA model implemented in the forecast package^22^ and the same filter was applied to the resistance series. Cross-correlations between the residual series were evaluated from -12 to +12 months. Negative lags indicated that prescribing preceded resistance; positive lags indicated that resistance preceded prescribing. The same prewhitened cross-correlation framework was applied to pairwise antibiotic-resistance series within each organism to identify temporally ordered resistance signals across antibiotics. Findings were interpreted as ecological temporal associations, not causal effects.

### Software

Analyses were conducted using Python 3.10.16 (Python Software Foundation; https://www.python.org) and R 4.1.3 (R Core Team; https://www.R-project.org). In Python, pandas^23^ (version 2.3.3) was used for data cleaning and aggregation, NumPy^24^ (2.2.6) for numerical calculations and bootstrap resampling, and Matplotlib^25^ (3.10.8) for figures. In R, data.table^26^ (1.15.4) and the tidyverse packages dplyr and tidyr^27^ (1.1.4 and 1.3.1, respectively) were used for data management, and zoo^28^ (1.8-12) for monthly time indices. Generalised linear mixed models were fitted with glmmTMB^20^ (1.1.7), and automated ARIMA selection and prewhitening used forecast^22^ (8.21).

### Patient consent and ethics approval

This study has been approved by the Ethics Committee of the Indraprastha Institute of Information Technology, Delhi(IRB-IIITD/FR/07/2025/01). This study utilized aggregated datasets routinely generated at TATA 1mg. Patient consent is covered under the privacy policy of TATA 1mg’s platform. No person was sampled specifically for this work.

## Role of the funding source

This work was supported by The Trinity Challenge grant titled AMRSense: Empowering Communities with a Proactive OneHealth Ecosystem. The funding agency had no role in the study design or analysis of this work.

## Data Availability

Individual or aggregated data from TATA 1mg is not publicly accessible. Requests for aggregated datasets should be directed to Nikhil Doegar, Senior Vice President, TATA 1mg and are subject to approval.

## Code Availability

All custom code used for data analysis is available at the GitHub repository https://github.com/tavlab-iiitd/SensingCommunityAMR

## Confict of Interest

VM, RS, ND and GA are employees of Tata 1mg Technologies Private Limited. ZA is a joint intern with IIITD and TATA 1mg. The remaining authors declare no competing interests.

## Author Contributions

TS, JK, GA, ND, NG, TB, SG, AM and HS conceptualised the research. VM performed data extraction and aggregation from the Tata 1mg database. JK performed data preprocessing and WHO AWaRe classification/matching. JK and VM conducted the formal analyses and prepared the visualisations. JK, VM, TS, RS, ND and GA interpreted the findings and contributed to drafting the manuscript. ZA, JK, TS and GA contributed to the independent platform-based survey and interpretation of its findings alongside the primary analysis. All authors critically reviewed and approved the final manuscript.

## Notes

### Author Declarations

This study has been approved by the Ethics Committee of the Indraprastha Institute of Information Technology, Delhi with approval number IRB-IIITD/FR/07/2025/01. This study utilized aggregated datasets routinely generated at TATA 1mg. Patient consent is covered under the privacy policy of TATA 1mg platform. No person was sampled specifically for this work.

### Summary of Updates

Updated the analyses and added comparisons with national AMR surveillance networks. Also, revised the Methods, Results, figures, Discussion and supplementary files to incorporate the changes and enhance clarity and contextualisation.

