## Supplementary tables for "Antibiotic prescribing and resistance beyond hospital surveillance networks in India"

**Supplementary Table 1. Scale, geographic coverage and AWaRe composition of diagnostic dataset from Tata 1mg, 2023-2025. (N = 155,764 samples analysed)**

| **Category** | **Subcategory** | **N** | **%** | **Notes** |
| --- | --- | --- | --- | --- |
| Geographic coverage | States represented | 23 | - |  |
|  | Districts represented | 102 | - |  |
|  | Pincodes represented | 1661 |  |  |
| Temporal coverage | Study period | 2023-2025 | - |  |
|  | Total months | 36 | - |  |
| Total possible state-month cells | States × Months | 828 |  |  |
| Observed state-month cells | Cells with any activity | 771 |  |  |
| Active state-month proportion (%) | Observed / possible | 93.12 |  |  |
| Culture outcome | Culture positive | 35,657 | 22.89 | a |
|  | No growth | 120107 | 77.11 | a |
| Sample type (overall) | Urine | 146273 | 93.91 | a |
|  | Blood | 5944 | 3.82 | a |
|  | Sputum | 3547 | 2.28 | a |
| Sample type (culture positive) | Urine | 34584 | 96.99 | b |
|  | Blood | 559 | 1.57 | b |
|  | Sputum | 514 | 1.44 | b |
| Top contributing states overall | Karnataka | 29161 | 18.72 | a |
|  | Delhi | 26466 | 16.99 | a |
|  | Haryana | 21421 | 13.75 | a |
|  | Uttar Pradesh | 20635 | 13.25 | a |
|  | Maharashtra | 20578 | 13.21 | a |
| Top contributing states for positive cultures | Karnataka | 9631 | 27.01 | b |
|  | Maharashtra | 7476 | 20.97 | b |
|  | Delhi | 4613 | 12.94 | b |
|  | Haryana | 3987 | 11.18 | b |
|  | Telangana | 3612 | 10.13 | b |
| Most frequently isolated organisms overall | *Escherichia coli* | 21608 | 60.6 | b |
|  | *Klebsiella pneumoniae* | 5997 | 16.8 | b |
|  | *Pseudomonas aeruginosa* | 1831 | 5.14 | b |
| Most frequently isolated organisms in Urine | *Escherichia coli* | 21549 | 62.31 | c |
|  | *Klebsiella pneumoniae* | 5759 | 16.65 | c |
|  | *Pseudomonas aeruginosa* | 1629 | 4.71 | c |
| Most frequently isolated organisms in Blood | *Salmonella Typhi* | 305 | 54.56 | d |
|  | *Salmonella Paratyphi* | 164 | 29.34 | d |
| Most frequently isolated organisms in Sputum | *Klebsiella pneumoniae* | 226 | 43.97 | e |
|  | *Pseudomonas aeruginosa* | 200 | 38.91 | e |

a Percentages calculated out of total samples analysed.

b Percentages calculated among culture-positive isolates.

c Percentages calculated among urine culture-positive isolates.

d Percentages calculated among blood culture-positive isolates.

e Percentages calculated among sputum culture-positive isolates.

**Supplementary Table 2. Antimicrobial resistance profiles by specimen type, organism and antibiotic.**

| **Aware** | **Antibiotic** | **URINE** | | | **BLOOD** | | **SPUTUM** | |
| --- | --- | --- | --- | --- | --- | --- | --- | --- |
|  |  | ***E. coli*** | ***K. pneumoniae*** | ***P. aeruginosa*** | ***S. typhi*** | ***S. paratyphi-A*** | ***K. pneumoniae*** | ***P. aeruginosa*** |
| Reserve | Aztreonam | 216/5902 (3.66%) | 382/2508 (15.23%) | 203/1461 (13.89%) | - | - | 4/153 (2.61%) | 3/200 (1.50%) |
|  | Colistin | 0/95 (0.00%) | 1/216 (0.46%) | 5/110 (4.55%) | - | - | 0/3* | 0/5* |
|  | Tigecycline | 55/5843 (0.94%) | 75/2451 (3.06%) | IR | - | - | 3/155 (1.94%) | IR |
| Watch | Azithromycin | - | - | - | 11/305 (3.61%) | 4/164 (2.44%) | - | - |
|  | Cefepime | 4267/10191 (41.87%) | 1322/4155 (31.82%) | 216/1537 (14.05%) | 1/305 (0.33%) | 0/164 (0.00%) | 26/224 (11.61%) | 4/200 (2.00%) |
|  | Cefixime | 7218/10244 (70.46%) | 1118/2278 (49.08%) | IR | 9/12* | 4/8* | - | - |
|  | Cefotaxime | 5141/11283 (45.56%) | 883/3478 (25.39%) | IR | - | - | 21/169 (12.43%) | IR |
|  | Cefoxitin | 2228/8102 (27.50%) | 612/2059 (29.72%) | - | - | - | 4/25* | - |
|  | Ceftazidime | 6764/16837 (40.17%) | 1330/4815 (27.62%) | 294/1627 (18.07%) | - | - | 19/143 (13.29%) | 6/200 (3.00%) |
|  | Ceftriaxone | 10499/19594 (53.58%) | 1853/5751 (32.22%) | IR | 14/305 (4.59%) | 5/164 (3.05%) | 28/223 (12.56%) | IR |
|  | Cefuroxime | 4387/9358 (46.88%) | 943/3484 (27.07%) | IR | - | - | 31/226 (13.72%) | IR |
|  | Ciprofloxacin | 12275/21542 (56.98%) | 1911/5755 (33.21%) | 368/1629 (22.59%) | 154/304 (50.66%) | 163/164 (99.39%) | 24/225 (10.67%) | 21/200 (10.50%) |
|  | Doripenem | - | - | 0/3* | - | - | - | - |
|  | Ertapenem | 417/11802 (3.53%) | 315/2016 (15.62%) | IR | 1/305 (0.33%) | 0/164 (0.00%) | 2/15* | - |
|  | Fosfomycin | 199/19431 (1.02%) | 9/166 (5.42%) | - | - | - | - | - |
|  | Imipenem | 220/11085 (1.98%) | 219/3484 (6.29%) | 243/1614 (15.06%) | 1/305 (0.33%) | 0/164 (0.00%) | 7/226 (3.10%) | 6/200 (3.00%) |
|  | Levofloxacin | 1979/3638 (54.40%) | 415/1384 (29.99%) | 364/1531 (23.78%) | - | - | 19/200 (9.50%) | 29/200 (14.50%) |
|  | Meropenem | 307/10635 (2.89%) | 313/3827 (8.18%) | 212/1537 (13.79%) | 1/305 (0.33%) | 1/164 (0.61%) | 6/224 (2.68%) | 5/200 (2.50%) |
|  | Nalidixic acid | 8682/11296 (76.86%) | 1116/3461 (32.25%) | - | - | - | 0/1* | - |
|  | Norfloxacin | 10966/21407 (51.23%) | 1554/5729 (27.13%) | 192/1168 (16.44%) | - | - | 0/2* | 0/1* |
|  | Ofloxacin | 7775/14789 (52.57%) | 1212/4550 (26.64%) | 112/787 (14.23%) | - | - | 2/47 (4.26%) | 0/15* |
|  | Piperacillin | 1328/3057 (43.44%) | 267/1251 (21.34%) | 8/389 (2.06%) | - | - | 8/77 (10.39%) | 1/78 (1.28%) |
|  | Piperacillin/Tazobactam | 2166/21176 (10.23%) | 985/5633 (17.49%) | 166/1601 (10.37%) | 1/305 (0.33%) | 0/164 (0.00%) | 7/225 (3.11%) | 3/200 (1.50%) |
|  | Ticarcillin | 6848/9030 (75.84%) | IR | 1/1* | - | - | - | - |
|  | Ticarcillin/Clavulanic acid | - | - | 1/5* | - | - | - | - |
|  | Tobramycin | 10/12* | 3/4* | 162/703 (23.04%) | - | - | - | 2/56 (3.57%) |
| Access | Amikacin | 1050/21521 (4.88%) | 727/5744 (12.66%) | 196/1625 (12.06%) | - | - | 4/226 (1.77%) | 4/200 (2.00%) |
|  | Amoxicillin/Clavulanic acid | 4999/21537 (23.21%) | 1313/5758 (22.80%) | IR | 1/305 (0.33%) | 0/164 (0.00%) | 18/226 (7.96%) | IR |
|  | Ampicillin | 16818/20311 (82.80%) | IR | IR | - | - | IR | IR |
|  | Chloramphenicol | - | - | - | 2/305 (0.66%) | 0/164 (0.00%) | 0/5* | - |
|  | Gentamicin | 2138/20341 (10.51%) | 695/5409 (12.85%) | 114/598 (19.06%) | - | - | 6/213 (2.82%) | 2/45 (4.44%) |
|  | Nitrofurantoin | 1060/21539 (4.92%) | 1382/5754 (24.02%) | IR | - | - | 0/2* | IR |
|  | Tetracycline | 2289/5878 (38.94%) | 528/2558 (20.64%) | IR | - | - | 12/118 (10.17%) | - |
|  | Trimethoprim/Sulfamethoxazole | 8705/21401 (40.68%) | 1588/5716 (27.78%) | IR | 3/305 (0.98%) | 0/164 (0.00%) | 23/226 (10.18%) | - |
| Not recommended | Cefoperazone/Sulbactam | 134/1200 (11.17%) | 96/345 (27.83%) | 103/429 (24.01%) | 1/305 (0.33%) | 1/164 (0.61%) | 3/17* | 2/22* |

IR- Intrinsically resistant; *- n below 30

**Supplementary Table 3. Characteristics of the antibiotic prescription dataset recorded through the Tata 1mg digital pharmacy platform 2023-2025 (Total medicines prescribed = 25735715)**

| **Category** | **Subcategory** | **N** | **%** |
| --- | --- | --- | --- |
| Total antibiotics prescribed | - | 2784503 | 10.82 |
| Geographic coverage | States and UTs represented | 35 | - |
|  | Districts represented | 694 | - |
|  | Pincodes represented | 13584 | - |
|  | States retained for state-specific analyses (>=1,000 prescriptions) | 25 | 99.77 |
| Temporal coverage | Study period | 2023-2025 | - |
| Median | Prescription units per state-month | 437 | - |
| IQR | Prescription units per state-month (Q1-Q3) | 18-3409 | - |
| Top contributing states | Maharashtra | 6,02,263 | 21.63 |
|  | Andhra Pradesh | 3,64,089 | 13.08 |
|  | Uttar Pradesh | 2,91,032 | 10.45 |
|  | Delhi | 2,33,573 | 8.39 |
|  | Karnataka | 1,85,652 | 6.67 |
| AWaRe classification | Access | 7,98,643 | 28.68 |
|  | Watch | 13,07,563 | 46.96 |
|  | Reserve | 93,562 | 3.36 |
|  | Not recommended | 3,31,992 | 11.92 |
|  | Unclassified | 2,52,743 | 9.08 |
| Antibiotic classes | Beta-lactam/beta-lactamase-inhibitor | 5,89,954 | 21.19 |
|  | Macrolides | 5,69,675 | 20.46 |
|  | Third-generation-cephalosporins | 4,32,635 | 15.54 |
|  | Not recommended FDC | 3,31,992 | 11.92 |
|  | Unclassified | 2,52,743 | 9.08 |
|  | Second-generation-cephalosporins | 1,86,530 | 6.70 |
|  | Tetracyclines | 89,611 | 3.22 |
|  | Lincosamides | 73,046 | 2.62 |
|  | Penicillins | 65,102 | 2.34 |
|  | Oxazolidinones | 55,239 | 1.98 |
|  | Fluoroquinolones | 46,283 | 1.66 |
|  | Penems | 37,857 | 1.36 |
|  | First-generation-cephalosporins | 28,286 | 1.02 |
|  | Aminoglycosides | 13,238 | 0.48 |
|  | Beta-lactam/beta-lactamase-inhibitor (anti-pseudomonal) | 6,026 | 0.22 |
|  | Carbapenems | 5,659 | 0.20 |
|  | Fourth-generation-cephalosporins | 324 | 0.01 |

National descriptive summaries included all antibiotic prescriptions. State-specific analyses were restricted to 25 states and union territories with at least 1,000 prescriptions (99.77% of the total). Ten states or union territories were below this threshold and were excluded only from state-specific analyses (2,742 prescriptions; 0.10%): Himachal Pradesh (807), Goa (511), Pondicherry (389), Meghalaya (305), Nagaland (257), Sikkim (138), Arunachal Pradesh (132), Mizoram (86), Andaman & Nicobar Islands (66), and Dadra & Nagar Haveli (51). A further 3,675 prescriptions (0.13%) without a recorded state were retained in national summaries but excluded from geographic and state-specific analyses.

Seventy-two medicines could not be mapped to an entry in the WHO 2025 AWaRe or Not Recommended lists and were classified as Unclassified.

Doxycycline/lactobacillus, Clindamycin/clotrimazole, Norfloxacin/lactobacillus, Amoxicillin/clavulanic acid/lactobacillus, Cefixime/azithromycin/lactobacillus, Cefixime/lactobacillus, Cefalexin/clavulanic acid, Cefixime/dicloxacillin/lactobacillus, Amoxicillin/cloxacillin/lactobacillus, Ofloxacin/flavoxate, Faropenem/clavulanic acid, Cefixime/cloxacillin/lactobacillus, Amoxicillin/dicloxacillin/lactobacillus, Amoxicillin/lactobacillus, Cefixime/sulbactam, Ambroxol/doxycycline, Cefpodoxime proxetil/cloxacillin/lactobacillus, Cefuroxime/linezolid, Cefpodoxime proxetil/lactobacillus/dicloxacillin, Roxithromycin/ambroxol, Balofloxacin, Cefixime/ofloxacin/lactobacillus, Azithromycin/lactic acid bacillus, Cefuroxime/lactobacillus, Doxycycline/bromelains, Doxycycline/beta-cyclodextrin, Clindamycin/disodium edetate, Ceftriaxone/disodium edetate/sulbactam, Ampicillin/dicloxacillin/lactobacillus, Levofloxacin/ambroxol, Amoxicillin/bromhexine, Ciprofloxacin/phenazopyridine, Azithromycin/adapalene, Cefpirome/sulbactam, Clindamycin/miconazole, Ampicillin/cloxacillin/lactobacillus, Cefpodoxime proxetil/lactobacillus, Zinc acetate/doxycycline/ivermectin, Amoxicillin/cloxacillin/lactobacillus/serrapeptase, Cefixime/acetylcysteine, Levofloxacin/lactobacillus, Amoxicillin/ambroxol, Cefadroxil/lactobacillus acidophilus, Ampicillin/lactobacillus, Amoxicillin/carbocisteine, Cefalexin/probenecid, Cefalexin/carbocisteine, Amoxicillin/lactobacillus/serrapeptase, Doxycycline/ornidazole/beta-cyclodextrin, Cefixime/clavulanic acid/lactobacillus, Meropenem/tazobactam, Cefixime/ornidazole/lactobacillus, Cefdinir/lactobacillus, Ampicillin/probenecid, Azithromycin/ambroxol, Cefadroxil/probenecid, Amoxicillin/tinidazole, Doxycycline/serrapeptase, Doxycycline/vitamin C, Cefpodoxime proxetil/ofloxacin/Bacillus coagulans, Cefixime/ambroxol/lactobacillus, Azithromycin/cefpodoxime proxetil/Saccharomyces boulardii, Amoxicillin/probenecid, Enrofloxacin, Cefixime/Saccharomyces boulardii/lactobacillus, Linezolid/sulbactam, Cefixime/linezolid/lactic acid bacillus, Amoxicillin/cloxacillin/probenecid, Atropine/tetracycline, Oxytetracycline/vitamin B6 (pyridoxine)/vitamin B3 (nicotinic acid)/vitamin B2/calcium/folic acid/methylcobalamin/thiamine (vitamin B1), Cefalexin/bromhexine, Ambroxol/cefadroxil.

**Supplementary Table 4: Descriptive pincode-level changes in antibiotic prescribing share and urinary antimicrobial resistance, 2023–2025. a) *E. coli***

| **Antibiotic** | **N₂** | **Both increased** | **Both decreased** | **Opposite directions** | **At least one unchanged** | **N₃** | **Successive annual increases in both** |
| --- | --- | --- | --- | --- | --- | --- | --- |
| Amikacin | 630 | 67 (10.6) | 35 (5.6) | 85 (13.5) | 443 (70.3) | 553 | 4 (0.7) |
| Amoxicillin/clavulanic acid | 631 | 149 (23.6) | 104 (16.5) | 249 (39.5) | 129 (20.4) | 553 | 12 (2.2) |
| Ampicillin | 614 | 26 (4.2) | 17 (2.8) | 41 (6.7) | 530 (86.3) | 544 | 2 (0.4) |
| Aztreonam | 97 | 1 (1.0) | 0 (0.0) | 2 (2.1) | 94 (96.9) | 89 | 0 (0.0) |
| Cefepime | 288 | 4 (1.4) | 3 (1.0) | 11 (3.8) | 270 (93.8) | 229 | 0 (0.0) |
| Cefixime | 281 | 61 (21.7) | 61 (21.7) | 111 (39.5) | 48 (17.1) | 238 | 9 (3.8) |
| Cefoperazone/sulbactam | 1 | 1 (100.0) | 0 (0.0) | 0 (0.0) | 0 (0.0) | 0 | — |
| Cefotaxime | 292 | 4 (1.4) | 11 (3.8) | 15 (5.1) | 262 (89.7) | 270 | 0 (0.0) |
| Ceftazidime | 345 | 6 (1.7) | 10 (2.9) | 14 (4.1) | 315 (91.3) | 311 | 0 (0.0) |
| Ceftriaxone | 550 | 68 (12.4) | 63 (11.5) | 155 (28.2) | 264 (48.0) | 478 | 5 (1.0) |
| Cefuroxime | 214 | 46 (21.5) | 57 (26.6) | 96 (44.9) | 15 (7.0) | 197 | 7 (3.6) |
| Ciprofloxacin | 630 | 6 (1.0) | 9 (1.4) | 16 (2.5) | 599 (95.1) | 553 | 0 (0.0) |
| Ertapenem | 357 | 20 (5.6) | 0 (0.0) | 17 (4.8) | 320 (89.6) | 310 | 1 (0.3) |
| Levofloxacin | 122 | 1 (0.8) | 3 (2.5) | 5 (4.1) | 113 (92.6) | 110 | 0 (0.0) |
| Meropenem | 261 | 15 (5.7) | 13 (5.0) | 26 (10.0) | 207 (79.3) | 215 | 0 (0.0) |
| Norfloxacin | 629 | 31 (4.9) | 53 (8.4) | 83 (13.2) | 462 (73.4) | 551 | 2 (0.4) |
| Ofloxacin | 282 | 46 (16.3) | 64 (22.7) | 110 (39.0) | 62 (22.0) | 248 | 4 (1.6) |
| Piperacillin | 43 | 0 (0.0) | 0 (0.0) | 1 (2.3) | 42 (97.7) | 43 | 0 (0.0) |
| Piperacillin/tazobactam | 630 | 65 (10.3) | 36 (5.7) | 93 (14.8) | 436 (69.2) | 548 | 6 (1.1) |
| Sulfamethoxazole/trimethoprim | 630 | 0 (0.0) | 0 (0.0) | 2 (0.3) | 628 (99.7) | 552 | 0 (0.0) |
| Tetracycline | 99 | 5 (5.1) | 9 (9.1) | 22 (22.2) | 63 (63.6) | 90 | 1 (1.1) |
| Tobramycin | 0 | — | — | — | — | 0 | — |
| **Total comparisons** | **7,626** | **622 (8.2)** | **548 (7.2)** | **1,154 (15.1)** | **5,302 (69.5)** | **6,682** | **53 (0.8)** |

**a) *K. pnemoniae***

| **Antibiotic** | **N₂** | **Both increased** | **Both decreased** | **Opposite directions** | **At least one unchanged** | **N₃** | **Successive annual increases in both** |
| --- | --- | --- | --- | --- | --- | --- | --- |
| Amikacin | 294 | 33 (11.2) | 31 (10.5) | 56 (19.0) | 174 (59.2) | 232 | 2 (0.9) |
| Amoxicillin/clavulanic acid | 294 | 45 (15.3) | 42 (14.3) | 113 (38.4) | 94 (32.0) | 232 | 5 (2.2) |
| Ampicillin† | 279 | 0 (0.0) | 3 (1.1) | 7 (2.5) | 269 (96.4) | 226 | 0 (0.0) |
| Aztreonam | 91 | 0 (0.0) | 0 (0.0) | 2 (2.2) | 89 (97.8) | 80 | 0 (0.0) |
| Cefepime | 200 | 5 (2.5) | 3 (1.5) | 8 (4.0) | 184 (92.0) | 158 | 0 (0.0) |
| Cefixime | 109 | 36 (33.0) | 17 (15.6) | 39 (35.8) | 17 (15.6) | 79 | 6 (7.6) |
| Cefoperazone/sulbactam | 0 | — | — | — | — | 0 | — |
| Cefotaxime | 155 | 1 (0.6) | 4 (2.6) | 13 (8.4) | 137 (88.4) | 137 | 0 (0.0) |
| Ceftazidime | 178 | 2 (1.1) | 5 (2.8) | 9 (5.1) | 162 (91.0) | 150 | 0 (0.0) |
| Ceftriaxone | 294 | 46 (15.6) | 38 (12.9) | 78 (26.5) | 132 (44.9) | 232 | 4 (1.7) |
| Cefuroxime | 155 | 25 (16.1) | 34 (21.9) | 54 (34.8) | 42 (27.1) | 137 | 1 (0.7) |
| Ciprofloxacin | 294 | 10 (3.4) | 7 (2.4) | 4 (1.4) | 273 (92.9) | 232 | 0 (0.0) |
| Ertapenem | 97 | 11 (11.3) | 2 (2.1) | 13 (13.4) | 71 (73.2) | 69 | 0 (0.0) |
| Levofloxacin | 85 | 1 (1.2) | 0 (0.0) | 3 (3.5) | 81 (95.3) | 72 | 0 (0.0) |
| Meropenem | 173 | 7 (4.0) | 12 (6.9) | 31 (17.9) | 123 (71.1) | 140 | 0 (0.0) |
| Norfloxacin | 292 | 19 (6.5) | 28 (9.6) | 40 (13.7) | 205 (70.2) | 230 | 2 (0.9) |
| Ofloxacin | 176 | 21 (11.9) | 42 (23.9) | 52 (29.5) | 61 (34.7) | 146 | 1 (0.7) |
| Piperacillin | 19 | 0 (0.0) | 0 (0.0) | 0 (0.0) | 19 (100.0) | 18 | 0 (0.0) |
| Piperacillin/tazobactam | 291 | 29 (10.0) | 23 (7.9) | 57 (19.6) | 182 (62.5) | 228 | 2 (0.9) |
| Sulfamethoxazole/trimethoprim | 294 | 0 (0.0) | 0 (0.0) | 1 (0.3) | 293 (99.7) | 232 | 0 (0.0) |
| Tetracycline | 97 | 8 (8.2) | 11 (11.3) | 24 (24.7) | 54 (55.7) | 83 | 2 (2.4) |
| Tobramycin | 0 | — | — | — | — | 0 | — |
| **Total comparisons** | **3,867** | **299 (7.7)** | **302 (7.8)** | **604 (15.6)** | **2,662 (68.8)** | **3,113** | **25 (0.8)** |

**N₂:** number of pincodes with prescribing share and resistance calculable in both 2023 and 2025 for that organism–antibiotic pair. The four endpoint-direction percentages use N₂.

**N₃:** number with both measures calculable in 2023, 2024 and 2025. Successive annual increases require **2023 < 2024 < 2025 in both measures**; percentages use N₃.

Prescribing share is the same-drug record count divided by all antibiotic records in that pincode-year. Resistance is resistant isolates divided by all interpretable susceptibility results. Annual percentages use summed counts.

“At least one unchanged” includes unchanged zero prescribing share or resistance. An absent same-drug entry within an observed prescription year represents zero **recorded activity**; missing prescription or testing denominators are not assigned zero.

Totals count **pincode–organism–antibiotic comparisons**, not unique pincodes or independent samples. Percentages may not sum to 100 because of rounding. “—” indicates no calculable comparison.

**Supplementary Table 5. Monthly resistance trends in *Escherichia coli* urine isolates, 2023-2025**

| **Antibiotic** | **Model** | **Observed  resistance  (%)** | **States /  UTs** | **Pin-  codes** | **State-  months** | **Average change  pp/month  (95% CI)** | **Adjusted p  change** | **OR/month  (95% CI)** | **Adjusted p  OR** |
| --- | --- | --- | --- | --- | --- | --- | --- | --- | --- |
| Ampicillin | BB-RS | 82.82 | 15 | 988 | 496 | -0.0776  (-0.1827, 0.0274) | 0.2067 | 0.9970  (0.9864, 1.0078) | 0.7728 |
| Cefuroxime | BB-RI | 46.76 | 4 | 407 | 134 | -0.2013  (-0.3164, -0.0862) | **0.0016** | 0.9918  (0.9871, 0.9965) | **0.0022** |
| Amoxicillin/Clavulanic acid | BB-RS | 23.19 | 15 | 1019 | 516 | +0.1717  (0.0778, 0.2656) | **0.0010** | 1.0049  (0.9945, 1.0154) | 0.5063 |
| Cefotaxime | BB-RI | 45.53 | 5 | 505 | 167 | -0.2350  (-0.3413, -0.1287) | **1.55E-4** | 0.9905  (0.9862, 0.9948) | **1.65E-4** |
| Ceftriaxone | BB-RI | 53.56 | 14 | 911 | 479 | -0.0520  (-0.1250, 0.0211) | 0.2145 | 0.9978  (0.9948, 1.0009) | 0.2648 |
| Nitrofurantoin | BB-RI | 4.92 | 15 | 1019 | 517 | +0.0610  (0.0205, 0.1016) | **0.0075** | 1.0119  (1.0039, 1.0199) | **0.0093** |
| Ciprofloxacin | BB-RS | 56.94 | 15 | 1019 | 517 | -0.0258  (-0.1111, 0.0595) | 0.6774 | 0.9992  (0.9947, 1.0037) | 0.8323 |
| Gentamicin | B-RS | 10.47 | 15 | 988 | 496 | -0.0642  (-0.1220, -0.0063) | **0.0447** | 0.9928  (0.9867, 0.9990) | **0.0412** |
| Ceftazidime | BB-RI | 43.50 | 5 | 578 | 171 | -0.2024  (-0.3061, -0.0987) | **4.58E-4** | 0.9917  (0.9874, 0.9959) | **5.46E-4** |
| Norfloxacin | BB-RI | 51.21 | 15 | 1019 | 517 | -0.0953  (-0.1737, -0.0169) | **0.0277** | 0.9961  (0.9929, 0.9993) | **0.0332** |
| Imipenem | BB-RI | 1.97 | 5 | 505 | 162 | +0.0003  (-0.0368, 0.0374) | 0.9869 | 1.0001  (0.9864, 1.0141) | 0.9869 |
| Ofloxacin | BB-RI | 47.45 | 4 | 479 | 139 | -0.1817  (-0.3113, -0.0522) | **0.0114** | 0.9927  (0.9875, 0.9979) | **0.0143** |
| Cefepime | BB-RI | 41.66 | 7 | 606 | 225 | -0.4223  (-0.6345, -0.2100) | **4.05E-4** | 0.9798  (0.9697, 0.9900) | **5.46E-4** |
| Meropenem | B-RI | 2.25 | 4 | 444 | 136 | -0.0010  (-0.0477, 0.0457) | 0.9869 | 0.9997  (0.9872, 1.0124) | 0.9869 |
| Amikacin | BB-RI | 4.86 | 15 | 1019 | 517 | +0.0966  (0.0510, 0.1423) | **2.33E-4** | 1.0173  (1.0090, 1.0256) | **2.64E-4** |
| Piperacillin/Tazobactam | BB-RS | 10.21 | 15 | 1016 | 515 | +0.1312  (0.0423, 0.2201) | **0.0080** | 1.0066  (0.9942, 1.0192) | 0.4453 |
| Fosfomycin | BB-RS | 1.02 | 14 | 909 | 479 | -0.0094  (-0.0426, 0.0239) | 0.6774 | 0.9996  (0.9601, 1.0408) | 0.9869 |
| Cefixime | B-RS | 70.52 | 11 | 550 | 368 | -0.0292  (-0.1446, 0.0861) | 0.6845 | 0.9985  (0.9925, 1.0046) | 0.7892 |
| Ertapenem | BB-RI | 3.50 | 12 | 647 | 404 | +0.3978  (0.3206, 0.4751) | **1.26E-22** | 1.2104  (1.1733, 1.2486) | **4.84E-32** |
| Trimethoprim/Sulfamethoxazole | BB-RS | 40.64 | 15 | 1019 | 517 | -0.1867  (-0.2771, -0.0963) | **2.69E-4** | 0.9922  (0.9876, 0.9967) | **0.0023** |
| Ticarcillin | BB-RI | 76.06 | 10 | 458 | 321 | -0.1227  (-0.2193, -0.0261) | **0.0223** | 0.9934  (0.9881, 0.9986) | **0.0283** |

Model: BB, beta-binomial; B, binomial; RI, state random intercept; RS, state random intercept and time slope. All models used a logit link and a fixed effect of month. UTs, union territories; pp, percentage points; OR, odds ratio; CI, confidence interval.

Average changes are conditional marginal derivatives, equally weighted across observed state-months and incorporating fitted state random effects. Positive values indicate increases. ORs are exponentiated fixed time coefficients. The 95% CIs use the delta method for average changes and exponentiated Wald limits for ORs.

Adjusted p-values test zero average change or zero fixed time coefficient, respectively. Benjamini-Hochberg adjustment was applied separately to each set of tests within this organism. CIs are unadjusted. Bold adjusted p-values are below 0.05. Observed resistance is pooled over included state-months; the denominator includes resistant, intermediate and sensitive isolates.

**Supplementary Table 6. Monthly resistance trends in *Klebsiella pneumoniae* urine isolates, 2023-2025**

| **Antibiotic** | **Model** | **Observed  resistance  (%)** | **States /  UTs** | **Pin-  codes** | **State-  months** | **Average change  pp/month  (95% CI)** | **Adjusted p  change** | **OR/month  (95% CI)** | **Adjusted p  OR** |
| --- | --- | --- | --- | --- | --- | --- | --- | --- | --- |
| Amoxicillin/Clavulanic acid | BB-RS | 22.22 | 6 | 539 | 214 | -0.1377  (-0.2900, 0.0145) | 0.1526 | 0.9761  (0.9407, 1.0128) | 0.4252 |
| Ceftriaxone | BB-RS | 31.87 | 6 | 539 | 214 | -0.0978  (-0.2740, 0.0785) | 0.3872 | 0.9947  (0.9850, 1.0045) | 0.4252 |
| Nitrofurantoin | B-RS | 23.05 | 6 | 539 | 214 | +0.3977  (0.2494, 0.5460) | **1.03E-6** | 1.0077  (0.9798, 1.0365) | 0.6755 |
| Ciprofloxacin | BB-RI | 32.85 | 6 | 539 | 214 | -0.0891  (-0.2591, 0.0809) | 0.3872 | 0.9961  (0.9886, 1.0036) | 0.4252 |
| Gentamicin | B-RI | 12.53 | 6 | 518 | 207 | -0.0643  (-0.1859, 0.0573) | 0.3872 | 0.9954  (0.9870, 1.0040) | 0.4252 |
| Norfloxacin | BB-RI | 26.88 | 6 | 539 | 214 | -0.1769  (-0.3336, -0.0202) | 0.0627 | 0.9915  (0.9841, 0.9990) | 0.0729 |
| Ofloxacin | BB-RI | 23.29 | 3 | 338 | 101 | -0.2997  (-0.5128, -0.0865) | **0.0164** | 0.9840  (0.9728, 0.9954) | **0.0204** |
| Cefepime | BB-RI | 31.05 | 6 | 450 | 183 | -0.4022  (-0.6618, -0.1426) | **0.0084** | 0.9739  (0.9574, 0.9908) | **0.0117** |
| Amikacin | B-RI | 12.30 | 6 | 538 | 214 | -0.0272  (-0.1375, 0.0831) | 0.6773 | 0.9980  (0.9898, 1.0062) | 0.6755 |
| Piperacillin/Tazobactam | BB-RI | 16.93 | 6 | 536 | 214 | +0.0383  (-0.1045, 0.1810) | 0.6773 | 1.0023  (0.9938, 1.0109) | 0.6755 |
| Cefixime | B-RS | 50.25 | 4 | 227 | 136 | -0.0375  (-0.2705, 0.1956) | 0.7526 | 0.9985  (0.9890, 1.0081) | 0.7571 |
| Ertapenem | BB-RI | 16.52 | 4 | 223 | 136 | +0.8829  (0.6240, 1.1417) | **3.25E-10** | 1.1230  (1.0830, 1.1645) | **5.32E-9** |
| Trimethoprim/Sulfamethoxazole | BB-RI | 27.81 | 6 | 538 | 214 | -0.0926  (-0.2489, 0.0636) | 0.3872 | 0.9956  (0.9882, 1.0030) | 0.4252 |
| Tetracycline | BB-RI | 20.65 | 4 | 216 | 120 | -0.6238  (-0.8654, -0.3822) | **1.96E-6** | 0.9628  (0.9485, 0.9773) | **4.80E-6** |

Model: BB, beta-binomial; B, binomial; RI, state random intercept; RS, state random intercept and time slope. All models used a logit link and a fixed effect of month. UTs, union territories; pp, percentage points; OR, odds ratio; CI, confidence interval.

Average changes are conditional marginal derivatives, equally weighted across observed state-months and incorporating fitted state random effects. Positive values indicate increases. ORs are exponentiated fixed time coefficients. The 95% CIs use the delta method for average changes and exponentiated Wald limits for ORs.

Adjusted p-values test zero average change or zero fixed time coefficient, respectively. Benjamini-Hochberg adjustment was applied separately to each set of tests within this organism. CIs are unadjusted. Bold adjusted p-values are below 0.05. Observed resistance is pooled over included state-months; the denominator includes resistant, intermediate and sensitive isolates.

**Supplementary Table 7. Monthly resistance trends in *Pseudomonas aeruginosa* urine isolates, 2023-2025**

| **Antibiotic** | **Model** | **Observed  resistance  (%)** | **States /  UTs** | **Pin-  codes** | **State-  months** | **Average change  pp/month  (95% CI)** | **Adjusted p  change** | **OR/month  (95% CI)** | **Adjusted p  OR** |
| --- | --- | --- | --- | --- | --- | --- | --- | --- | --- |
| Levofloxacin | BB-RI | 22.80 | 5 | 332 | 166 | -0.1765  (-0.4640, 0.1110) | 0.4120 | 0.9911  (0.9768, 1.0056) | 0.4072 |
| Ciprofloxacin | BB-RI | 22.26 | 5 | 334 | 166 | -0.1786  (-0.4659, 0.1086) | 0.4120 | 0.9908  (0.9762, 1.0055) | 0.4072 |
| Ceftazidime | BB-RS | 16.86 | 5 | 334 | 166 | -0.2130  (-0.5049, 0.0790) | 0.4120 | 0.9795  (0.9528, 1.0069) | 0.4072 |
| Imipenem | BB-RS | 13.97 | 5 | 334 | 165 | +0.1106  (-0.1791, 0.4003) | 0.6816 | 0.9910  (0.9463, 1.0379) | 0.7901 |
| Cefepime | BB-RS | 13.35 | 5 | 334 | 166 | -0.1723  (-0.4438, 0.0991) | 0.4120 | 0.9783  (0.9476, 1.0099) | 0.4072 |
| Aztreonam | B-RS | 13.53 | 5 | 326 | 166 | +0.0268  (-0.2577, 0.3113) | 0.9603 | 0.9845  (0.9439, 1.0269) | 0.7018 |
| Meropenem | BB-RS | 12.78 | 5 | 334 | 166 | +0.0918  (-0.1957, 0.3793) | 0.6835 | 0.9895  (0.9449, 1.0362) | 0.7901 |
| Amikacin | BB-RI | 11.33 | 5 | 334 | 166 | +0.0034  (-0.2409, 0.2478) | 0.9781 | 1.0003  (0.9810, 1.0199) | 0.9781 |
| Piperacillin/Tazobactam | BB-RI | 10.23 | 5 | 332 | 164 | +0.1941  (-0.0208, 0.4090) | 0.4120 | 1.0160  (0.9974, 1.0350) | 0.4072 |

Model: BB, beta-binomial; B, binomial; RI, state random intercept; RS, state random intercept and time slope. All models used a logit link and a fixed effect of month. UTs, union territories; pp, percentage points; OR, odds ratio; CI, confidence interval.

Average changes are conditional marginal derivatives, equally weighted across observed state-months and incorporating fitted state random effects. Positive values indicate increases. ORs are exponentiated fixed time coefficients. The 95% CIs use the delta method for average changes and exponentiated Wald limits for ORs.

Adjusted p-values test zero average change or zero fixed time coefficient, respectively. Benjamini-Hochberg adjustment was applied separately to each set of tests within this organism. CIs are unadjusted. Bold adjusted p-values are below 0.05. Observed resistance is pooled over included state-months; the denominator includes resistant, intermediate and sensitive isolates.

**Supplementary Table 8. Monthly trends in antibiotic prescribing share among all antibiotic prescriptions, 2023-2025**

| **Antibiotic** | **AWaRe** | **Model** | **States /  UTs** | **State-  months** | **Mean  share  (%)** | **Average change  pp/month  (95% CI)** | **Adjusted p  change** | **OR/month  (95% CI)** | **Adjusted p  OR** |
| --- | --- | --- | --- | --- | --- | --- | --- | --- | --- |
| Amoxicillin/clavulanic acid | A | BB-RS | 25 | 900 | 20.295 | -0.008370  (-0.035486, 0.018747) | 0.6058 | 1.0000  (0.9953, 1.0048) | 0.9962 |
| Clindamycin | A | BB-RS | 25 | 900 | 3.251 | +0.063254  (0.053407, 0.073102) | **2.41E-35** | 1.0227  (1.0144, 1.0310) | **5.52E-7** |
| Amoxicillin | A | BB-RS | 25 | 900 | 1.669 | +0.001374  (-0.005043, 0.007792) | 0.6747 | 1.0052  (0.9984, 1.0120) | 0.2688 |
| Doxycycline | A | BB-RS | 25 | 900 | 1.069 | -0.003461  (-0.008047, 0.001125) | 0.1739 | 0.9974  (0.9838, 1.0111) | 0.8853 |
| Cefalexin | A | BB-RS | 25 | 900 | 0.675 | -0.003113  (-0.006129, -0.000097) | 0.0718 | 0.9962  (0.9869, 1.0056) | 0.6165 |
| Cefadroxil | A | BB-RS | 25 | 900 | 0.523 | -0.001842  (-0.003996, 0.000312) | 0.1339 | 1.0003  (0.9925, 1.0082) | 0.9962 |
| Benzylpenicillin | A | BB-RS | 25 | 900 | 0.500 | -0.003541  (-0.006458, -0.000625) | **0.0346** | 0.9911  (0.9827, 0.9997) | 0.1036 |
| Amikacin | A | BB-RS | 25 | 900 | 0.348 | -0.003738  (-0.005177, -0.002298) | **1.80E-6** | 0.9905  (0.9827, 0.9984) | 0.0609 |
| Sultamicillin | A | BB-RS | 25 | 900 | 0.207 | +0.002031  (0.000816, 0.003246) | **0.0026** | 1.0046  (0.9949, 1.0143) | 0.5970 |
| Tetracycline | A | BB-RS | 25 | 900 | 0.127 | +0.002104  (0.001008, 0.003200) | **5.62E-4** | 1.0249  (1.0085, 1.0416) | **0.0137** |
| Cefpodoxime proxetil/clavulanic acid | NR | BB-RS | 25 | 900 | 4.615 | +0.023378  (0.012570, 0.034185) | **1.12E-4** | 1.0067  (0.9982, 1.0152) | 0.2443 |
| Cefixime/ofloxacin | NR | BB-RS | 25 | 900 | 1.730 | -0.008273  (-0.015022, -0.001524) | **0.0384** | 0.9941  (0.9861, 1.0021) | 0.2443 |
| Cefuroxime/clavulanic acid | NR | BB-RS | 25 | 900 | 1.599 | +0.005302  (-0.000144, 0.010747) | 0.0940 | 1.0024  (0.9936, 1.0112) | 0.7461 |
| Cefixime/clavulanic acid | NR | BB-RS | 25 | 900 | 1.196 | -0.010550  (-0.015011, -0.006090) | **3.56E-5** | 0.9928  (0.9840, 1.0017) | 0.2443 |
| Cefpodoxime proxetil/ofloxacin | NR | BB-RS | 25 | 900 | 0.697 | -0.006172  (-0.009197, -0.003147) | **2.12E-4** | 0.9957  (0.9870, 1.0046) | 0.4938 |
| Cefoperazone/sulbactam | NR | BB-RS | 25 | 900 | 0.374 | -0.001085  (-0.002406, 0.000237) | 0.1538 | 0.9858  (0.9768, 0.9950) | **0.0244** |
| Ceftriaxone/sulbactam | NR | BB-RS | 25 | 900 | 0.159 | +0.000602  (-0.000482, 0.001686) | 0.3070 | 0.9866  (0.9746, 0.9987) | 0.1020 |
| Ampicillin/cloxacillin | NR | BB-RS | 25 | 900 | 0.150 | -0.000444  (-0.001388, 0.000500) | 0.3565 | 1.0008  (0.9930, 1.0087) | 0.8365 |
| Azithromycin/cefixime | NR | BB-RS | 25 | 900 | 0.071 | +0.000419  (-0.000311, 0.001149) | 0.3070 | 1.0020  (0.9831, 1.0213) | 0.8365 |
| Amoxicillin/dicloxacillin | NR | BB-RI | 25 | 900 | 0.058 | -0.001156  (-0.002124, -0.000188) | **0.0384** | 0.9808  (0.9659, 0.9959) | 0.0639 |
| Linezolid | R | BB-RS | 25 | 900 | 1.861 | -0.002847  (-0.008466, 0.002773) | 0.3680 | 1.0002  (0.9939, 1.0066) | 0.9424 |
| Faropenem | R | BB-RS | 25 | 900 | 1.482 | -0.002483  (-0.007890, 0.002923) | 0.3680 | 1.0057  (0.9960, 1.0156) | 0.5008 |
| Ceftazidime/avibactam | R | BB-RS | 25 | 900 | 0.011 | +0.000239  (0.000059, 0.000420) | **0.0185** | 1.0074  (0.9848, 1.0306) | 0.6984 |
| Aztreonam | R | BB-RI | 25 | 900 | 0.0054 | +0.000224  (0.000111, 0.000336) | **3.98E-4** | 1.0410  (1.0228, 1.0596) | **3.21E-5** |
| Azithromycin | W | BB-RS | 25 | 900 | 19.135 | -0.073310  (-0.097849, -0.048770) | **2.38E-8** | 0.9967  (0.9914, 1.0021) | 0.5725 |
| Cefixime | W | BB-RS | 25 | 900 | 7.109 | +0.009002  (-0.005844, 0.023848) | 0.2607 | 1.0037  (0.9953, 1.0121) | 0.6528 |
| Cefuroxime | W | BB-RS | 25 | 900 | 6.390 | -0.030543  (-0.045664, -0.015423) | **2.51E-4** | 1.0020  (0.9928, 1.0113) | 0.8237 |
| Cefpodoxime proxetil | W | BB-RS | 25 | 900 | 4.995 | +0.009621  (-0.002496, 0.021737) | 0.1709 | 1.0016  (0.9957, 1.0076) | 0.8237 |
| Minocycline | W | BB-RS | 25 | 900 | 1.935 | +0.024950  (0.018534, 0.031366) | **2.49E-13** | 1.0111  (1.0029, 1.0193) | **0.0411** |
| Clarithromycin | W | BB-RS | 25 | 900 | 1.875 | -0.007959  (-0.013674, -0.002244) | **0.0106** | 0.9962  (0.9894, 1.0032) | 0.5725 |
| Ofloxacin | W | BB-RS | 25 | 900 | 1.569 | -0.000085  (-0.004757, 0.004587) | 0.9715 | 0.9990  (0.9932, 1.0048) | 0.8237 |
| Lymecycline | W | BB-RS | 25 | 900 | 0.530 | +0.003859  (0.001334, 0.006385) | **0.0055** | 1.0079  (1.0018, 1.0141) | **0.0411** |
| Roxithromycin | W | BB-RS | 25 | 900 | 0.525 | -0.004065  (-0.006719, -0.001412) | **0.0055** | 0.9894  (0.9812, 0.9977) | **0.0411** |
| Ceftriaxone | W | BB-RS | 25 | 900 | 0.406 | -0.001198  (-0.003065, 0.000669) | 0.2606 | 1.0000  (0.9897, 1.0104) | 0.9984 |

Model: BB, beta-binomial; B, binomial; RI, state random intercept; RS, state random intercept and time slope. All models used a logit link and a fixed effect of month. UTs, union territories; pp, percentage points; OR, odds ratio; CI, confidence interval. AWaRe: A, Access; W, Watch; R, Reserve; NR, Not Recommended.

Average changes are conditional marginal derivatives, equally weighted across observed state-months and incorporating fitted state random effects. Positive values indicate increases. ORs are exponentiated fixed time coefficients. The 95% CIs use the delta method for average changes and exponentiated Wald limits for ORs.

Adjusted p-values test zero average change or zero fixed time coefficient, respectively. Benjamini-Hochberg adjustment was applied separately to each set of tests within AWaRe category for this denominator. CIs are unadjusted. Bold adjusted p-values are below 0.05. Mean share is the unweighted mean of observed state-month prescribing shares.

**Supplementary Table 9. Monthly trends in antibiotic prescribing share among all medicine prescriptions, 2023-2025**

| **Antibiotic** | **AWaRe** | **Model** | **States /  UTs** | **State-  months** | **Mean  share  (%)** | **Average change  pp/month  (95% CI)** | **Adjusted p  change** | **OR/month  (95% CI)** | **Adjusted p  OR** |
| --- | --- | --- | --- | --- | --- | --- | --- | --- | --- |
| Amoxicillin/clavulanic acid | A | BB-RS | 25 | 900 | 1.928 | -0.004069  (-0.006951, -0.001188) | **0.0063** | 0.9973  (0.9938, 1.0007) | 0.2235 |
| Clindamycin | A | BB-RS | 25 | 900 | 0.276 | +0.003879  (0.003247, 0.004510) | **2.25E-32** | 1.0183  (1.0102, 1.0265) | **8.96E-5** |
| Amoxicillin | A | BB-RS | 25 | 900 | 0.147 | +0.000217  (-0.000264, 0.000698) | 0.3770 | 1.0022  (0.9964, 1.0081) | 0.5088 |
| Doxycycline | A | BB-RS | 25 | 900 | 0.105 | -0.000795  (-0.001183, -0.000408) | **1.16E-4** | 0.9939  (0.9808, 1.0071) | 0.5088 |
| Cefalexin | A | BB-RS | 25 | 900 | 0.063 | -0.000631  (-0.000882, -0.000381) | **2.63E-6** | 0.9930  (0.9839, 1.0022) | 0.2235 |
| Cefadroxil | A | BB-RS | 25 | 900 | 0.052 | -0.000310  (-0.000501, -0.000119) | **0.0021** | 0.9966  (0.9883, 1.0050) | 0.5088 |
| Benzylpenicillin | A | BB-RS | 25 | 900 | 0.041 | -0.000525  (-0.000740, -0.000309) | **4.47E-6** | 0.9872  (0.9793, 0.9952) | **0.0090** |
| Amikacin | A | BB-RS | 25 | 900 | 0.036 | -0.000469  (-0.000611, -0.000327) | **4.88E-10** | 0.9883  (0.9804, 0.9963) | **0.0143** |
| Sultamicillin | A | BB-RS | 25 | 900 | 0.021 | +0.000170  (0.000064, 0.000275) | **0.0021** | 0.9995  (0.9896, 1.0095) | 0.9249 |
| Tetracycline | A | BB-RS | 25 | 900 | 0.011 | +0.000153  (0.000068, 0.000238) | **6.82E-4** | 1.0207  (1.0050, 1.0366) | **0.0237** |
| Cefpodoxime proxetil/clavulanic acid | NR | BB-RS | 25 | 900 | 0.462 | +0.000781  (-0.000380, 0.001942) | 0.2413 | 1.0022  (0.9932, 1.0113) | 0.8355 |
| Cefixime/ofloxacin | NR | BB-RS | 25 | 900 | 0.184 | -0.001309  (-0.002015, -0.000604) | **9.18E-4** | 0.9921  (0.9847, 0.9995) | 0.0933 |
| Cefuroxime/clavulanic acid | NR | BB-RS | 25 | 900 | 0.154 | +0.000086  (-0.000407, 0.000579) | 0.7318 | 0.9986  (0.9901, 1.0072) | 0.8355 |
| Cefixime/clavulanic acid | NR | BB-RS | 25 | 900 | 0.128 | -0.001966  (-0.002444, -0.001487) | **8.07E-15** | 0.9897  (0.9794, 1.0001) | 0.1061 |
| Cefpodoxime proxetil/ofloxacin | NR | BB-RS | 25 | 900 | 0.066 | -0.000634  (-0.000948, -0.000320) | **3.76E-4** | 0.9939  (0.9844, 1.0035) | 0.3523 |
| Cefoperazone/sulbactam | NR | BB-RS | 25 | 900 | 0.028 | -0.000168  (-0.000290, -0.000046) | **0.0174** | 0.9809  (0.9708, 0.9911) | **0.0025** |
| Ceftriaxone/sulbactam | NR | BB-RS | 25 | 900 | 0.015 | +0.000049  (-0.000048, 0.000146) | 0.3608 | 0.9832  (0.9717, 0.9947) | **0.0220** |
| Ampicillin/cloxacillin | NR | BB-RS | 25 | 900 | 0.013 | -0.000064  (-0.000136, 0.000008) | 0.1358 | 0.9985  (0.9907, 1.0063) | 0.8355 |
| Azithromycin/cefixime | NR | BB-RS | 25 | 900 | 0.011 | +0.000074  (-0.000038, 0.000187) | 0.2413 | 1.0006  (0.9815, 1.0201) | 0.9480 |
| Amoxicillin/dicloxacillin | NR | BB-RI | 25 | 900 | 0.0070 | -0.000142  (-0.000257, -0.000028) | **0.0299** | 0.9801  (0.9654, 0.9951) | **0.0309** |
| Linezolid | R | BB-RS | 25 | 900 | 0.179 | -0.000755  (-0.001270, -0.000241) | **0.0053** | 0.9968  (0.9900, 1.0037) | 0.7315 |
| Faropenem | R | BB-RS | 25 | 900 | 0.142 | -0.001343  (-0.001835, -0.000852) | **3.34E-7** | 1.0026  (0.9937, 1.0115) | 0.7591 |
| Ceftazidime/avibactam | R | BB-RS | 25 | 900 | 0.0011 | +0.000017  (0.000001, 0.000034) | **0.0370** | 1.0035  (0.9812, 1.0263) | 0.7593 |
| Aztreonam | R | BB-RI | 25 | 900 | 0.0005 | +0.000022  (0.000010, 0.000033) | **3.18E-4** | 1.0370  (1.0192, 1.0550) | **1.49E-4** |
| Azithromycin | W | BB-RS | 25 | 900 | 1.764 | -0.008772  (-0.011891, -0.005654) | **1.17E-7** | 0.9950  (0.9912, 0.9988) | 0.0541 |
| Cefixime | W | BB-RS | 25 | 900 | 0.717 | -0.002503  (-0.004064, -0.000942) | **0.0028** | 1.0003  (0.9902, 1.0104) | 0.9581 |
| Cefuroxime | W | BB-RS | 25 | 900 | 0.685 | -0.009182  (-0.011290, -0.007074) | **1.37E-16** | 0.9995  (0.9888, 1.0103) | 0.9581 |
| Cefpodoxime proxetil | W | BB-RS | 25 | 900 | 0.542 | +0.000714  (-0.000789, 0.002216) | 0.3910 | 0.9988  (0.9926, 1.0051) | 0.8938 |
| Clarithromycin | W | BB-RS | 25 | 900 | 0.182 | -0.001172  (-0.001741, -0.000604) | **1.07E-4** | 0.9924  (0.9852, 0.9997) | 0.1361 |
| Minocycline | W | BB-RS | 25 | 900 | 0.176 | +0.001147  (0.000661, 0.001634) | **9.45E-6** | 1.0065  (0.9985, 1.0146) | 0.2817 |
| Ofloxacin | W | BB-RS | 25 | 900 | 0.148 | -0.000552  (-0.000965, -0.000138) | **0.0128** | 0.9957  (0.9888, 1.0027) | 0.3861 |
| Lymecycline | W | BB-RS | 25 | 900 | 0.047 | +0.000060  (-0.000114, 0.000234) | 0.4967 | 1.0043  (0.9984, 1.0102) | 0.3104 |
| Roxithromycin | W | BB-RS | 25 | 900 | 0.047 | -0.000707  (-0.000895, -0.000519) | **8.82E-13** | 0.9855  (0.9793, 0.9918) | **6.29E-5** |
| Ceftriaxone | W | BB-RS | 25 | 900 | 0.043 | -0.000242  (-0.000436, -0.000047) | **0.0187** | 0.9969  (0.9870, 1.0069) | 0.7673 |

Model: BB, beta-binomial; B, binomial; RI, state random intercept; RS, state random intercept and time slope. All models used a logit link and a fixed effect of month. UTs, union territories; pp, percentage points; OR, odds ratio; CI, confidence interval. AWaRe: A, Access; W, Watch; R, Reserve; NR, Not Recommended.

Average changes are conditional marginal derivatives, equally weighted across observed state-months and incorporating fitted state random effects. Positive values indicate increases. ORs are exponentiated fixed time coefficients. The 95% CIs use the delta method for average changes and exponentiated Wald limits for ORs.

Adjusted p-values test zero average change or zero fixed time coefficient, respectively. Benjamini-Hochberg adjustment was applied separately to each set of tests within AWaRe category for this denominator. CIs are unadjusted. Bold adjusted p-values are below 0.05. Mean share is the unweighted mean of observed state-month prescribing shares.
