## Supplementary Figures for "Antibiotic prescribing and resistance beyond hospital surveillance networks in India"

a.
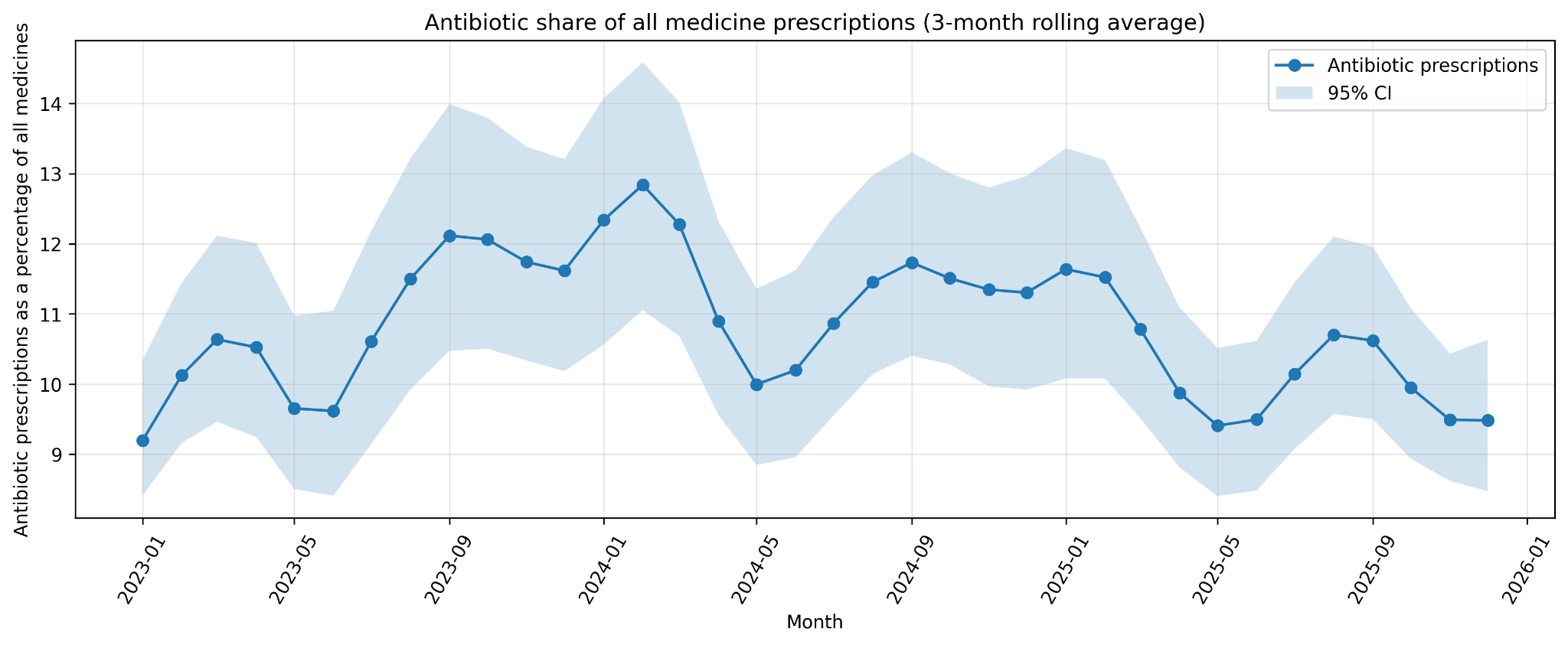

b.
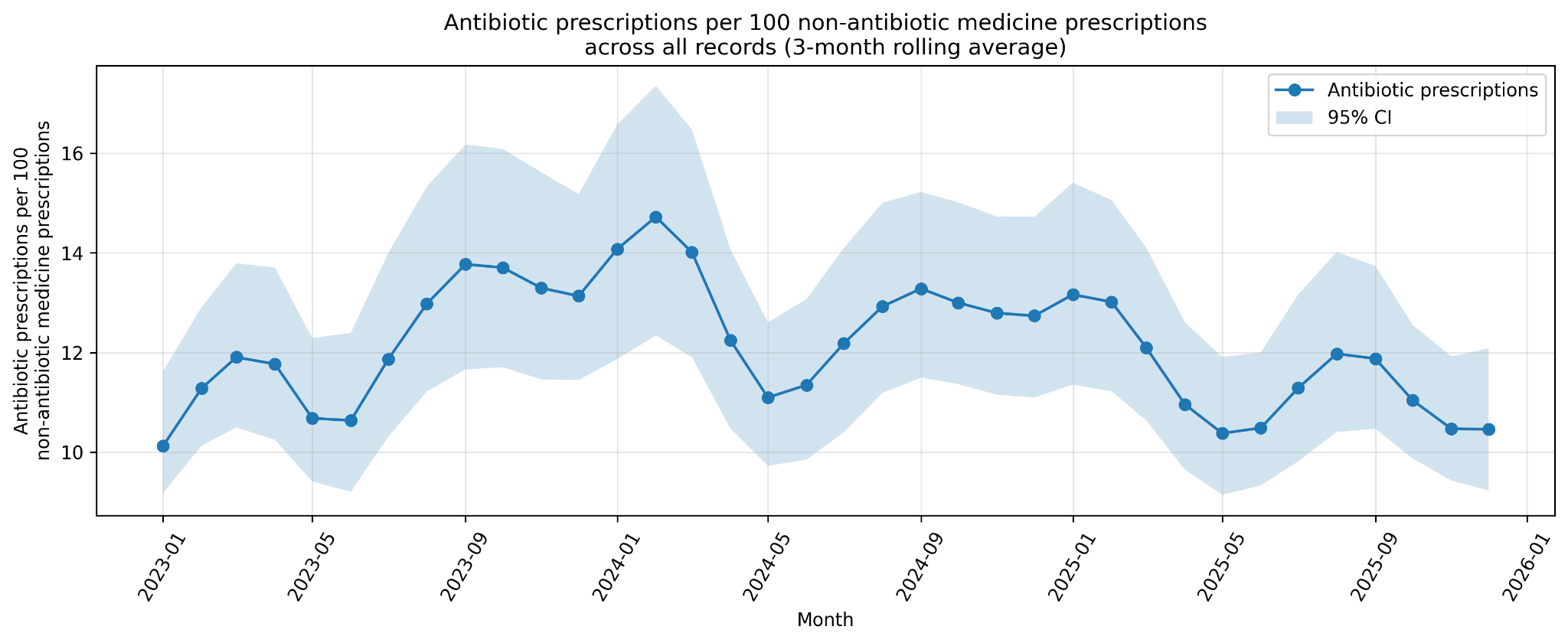

**Supplementary Figure 1: Temporal trend in total antibiotic share of digitised prescriptions, India, 2023–2025. a.** Trends in monthly antibiotic prescriptions are shown as the percentage of total platform medicine prescriptions **b.** Trend of antibiotic prescriptions as per 100 non antibiotic medicine prescriptions.

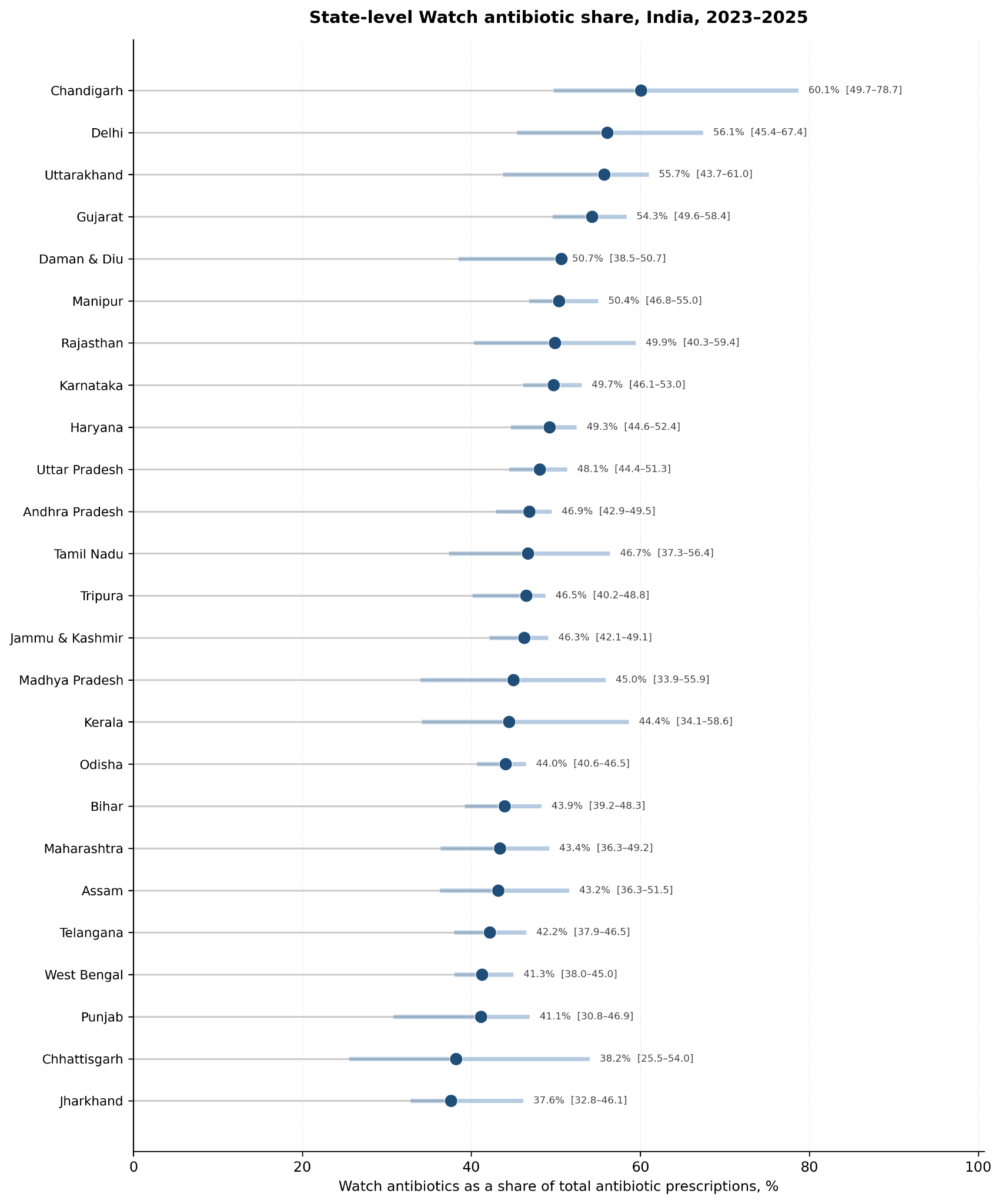

**Supplementary Figure 2: State/UT-level variation in Watch antibiotic prescriptions across the digital pharmacy platform, India, 2023-2025. Estimates are restricted to the 25 states/UTs contributing at least 1,000 antibiotic prescriptions.**

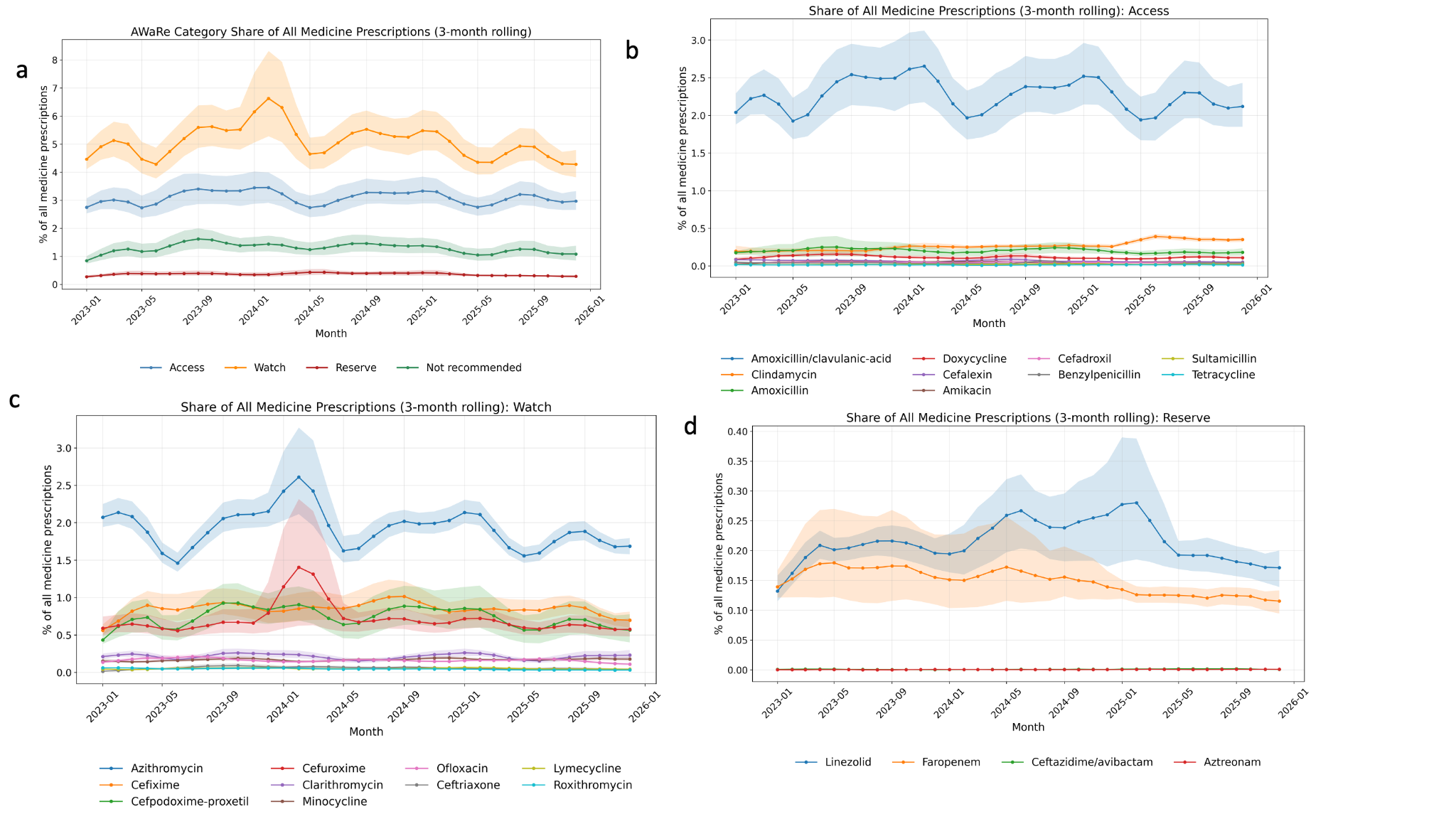

**Supplementary Figure 3. Temporal trends in antibiotic prescriptions from all medicines 2023–2025.**

Trends of antibiotic prescriptions are shown for (a) AWaRe category, (b) Access antibiotics, (c) Watch antibiotics, and (d) Reserve antibiotics

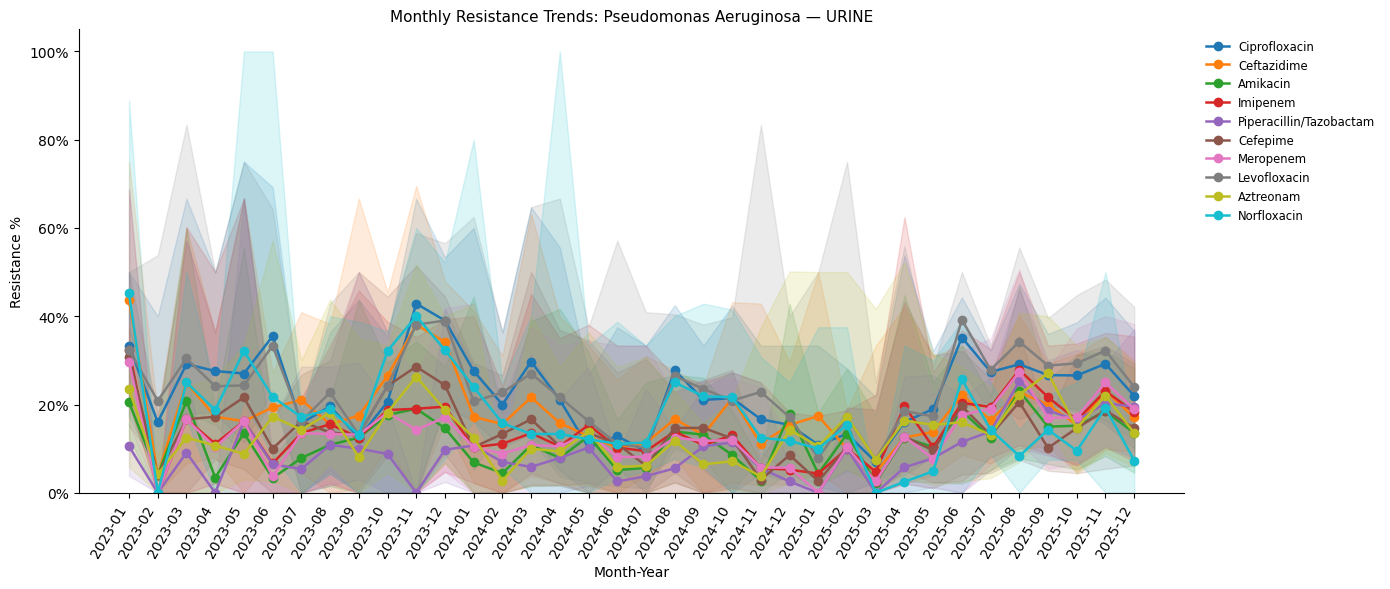

**Supplementary Figure 4. Bootstrapped temporal trends in antibiotic resistance for *P. aeruginosa* from Urine samples, 2023–2025.**

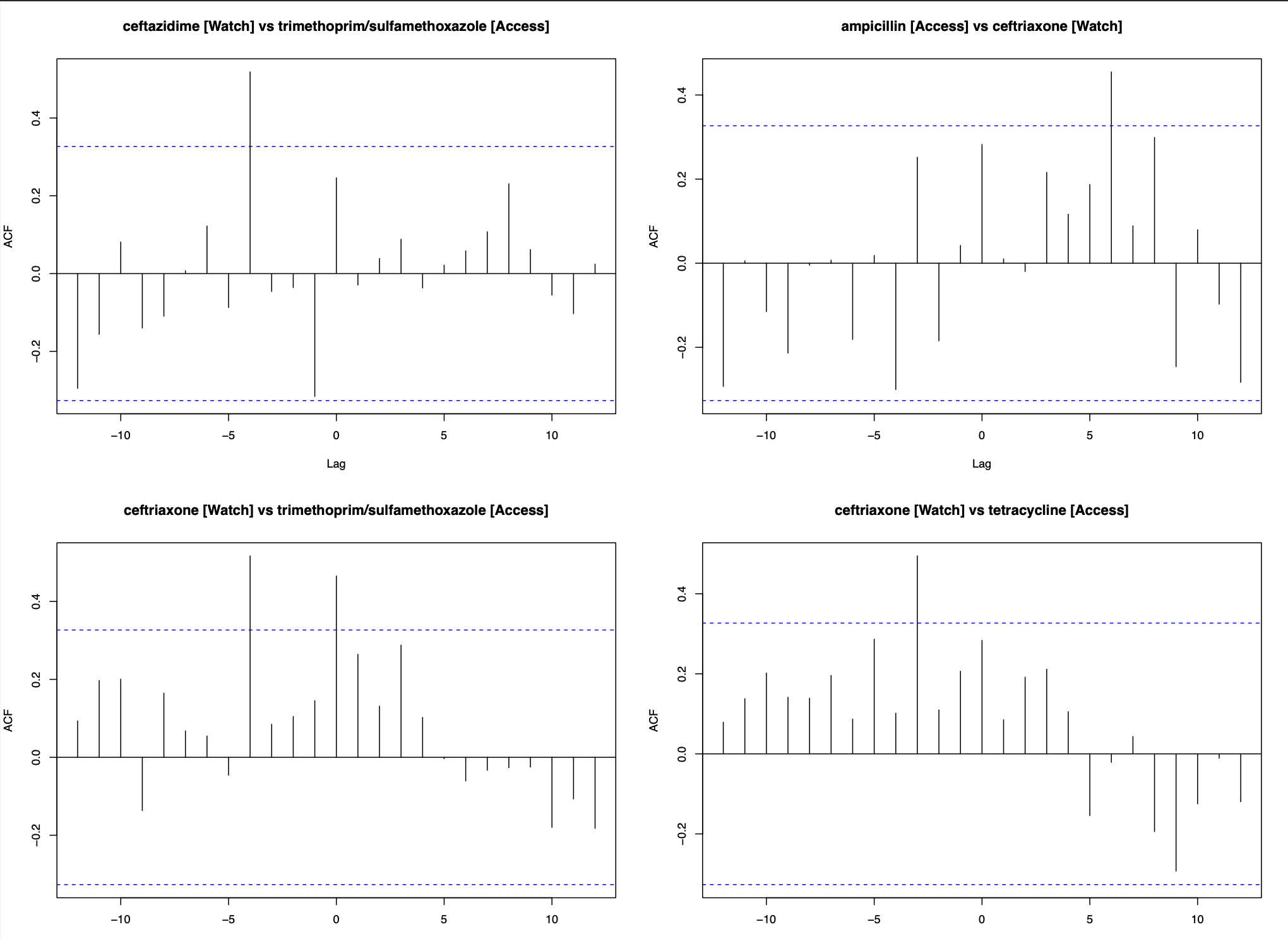

**Supplementary Figure 5. Pre-whitened cross-correlation plots between antibiotic series showing Watch-to-Access resistance lead–lag relationships in urinary *E. coli*.** For panels labelled A vs B, negative-lag peaks indicate that resistance to A preceded resistance to B; for Access-vs-Watch panels, positive-lag peaks indicate that the Watch-class comparator preceded the Access-class comparator. Significant threshold crossings show that resistance to Watch antibiotics, including ceftazidime and ceftriaxone, preceded resistance to Access antibiotics, including trimethoprim-sulfamethoxazole, ampicillin, and tetracycline by approximately 3–6 months. Dashed lines indicate the statistical significance threshold.

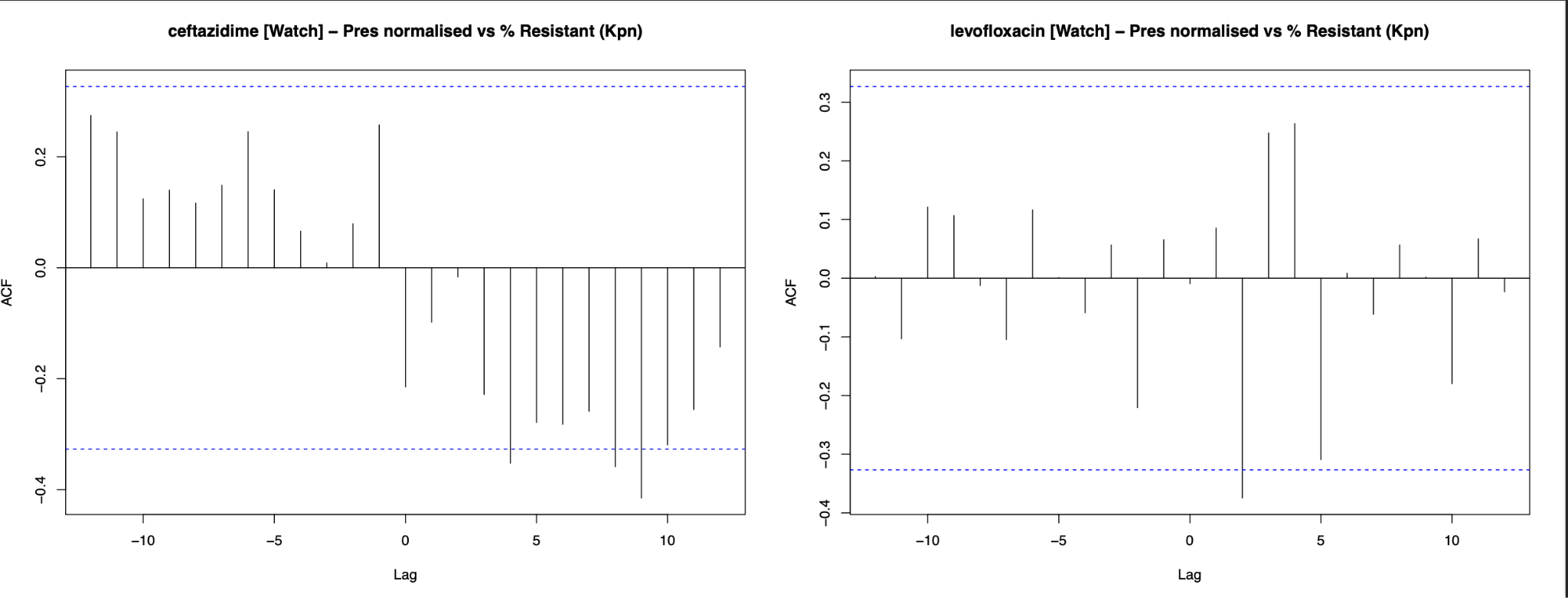

**Supplementary Figure 6: Pre-whitened cross-correlation between normalised ceftazidime and levofloxacin prescriptions and urinary *K. pneumoniae* resistance.** For panels labelled A vs B, negative-lag peaks indicate that A preceded B. The plot shows the cross-correlation function between normalised prescriptions and the percentage of antibiotic-resistant urinary *K. pneumoniae* isolates.

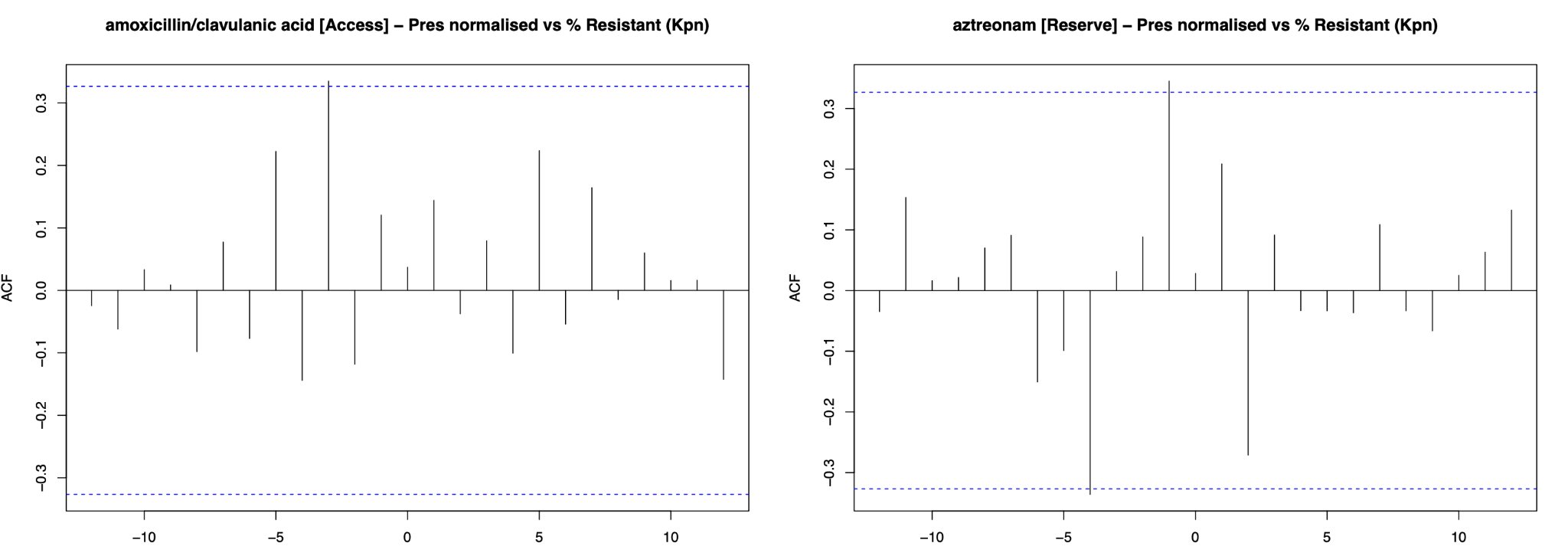

**Supplementary Figure 7: Pre-whitened cross-correlation between normalised amoxicillin-clavulanic acid and aztreonam prescriptions and urinary *K. pneumoniae* resistance.** For panels labelled A vs B, negative-lag peaks indicate that A preceded B. The plot shows the cross-correlation function between monthly normalised prescriptions and the monthly percentage of antibiotic-resistant urinary K. pneumoniae isolates.

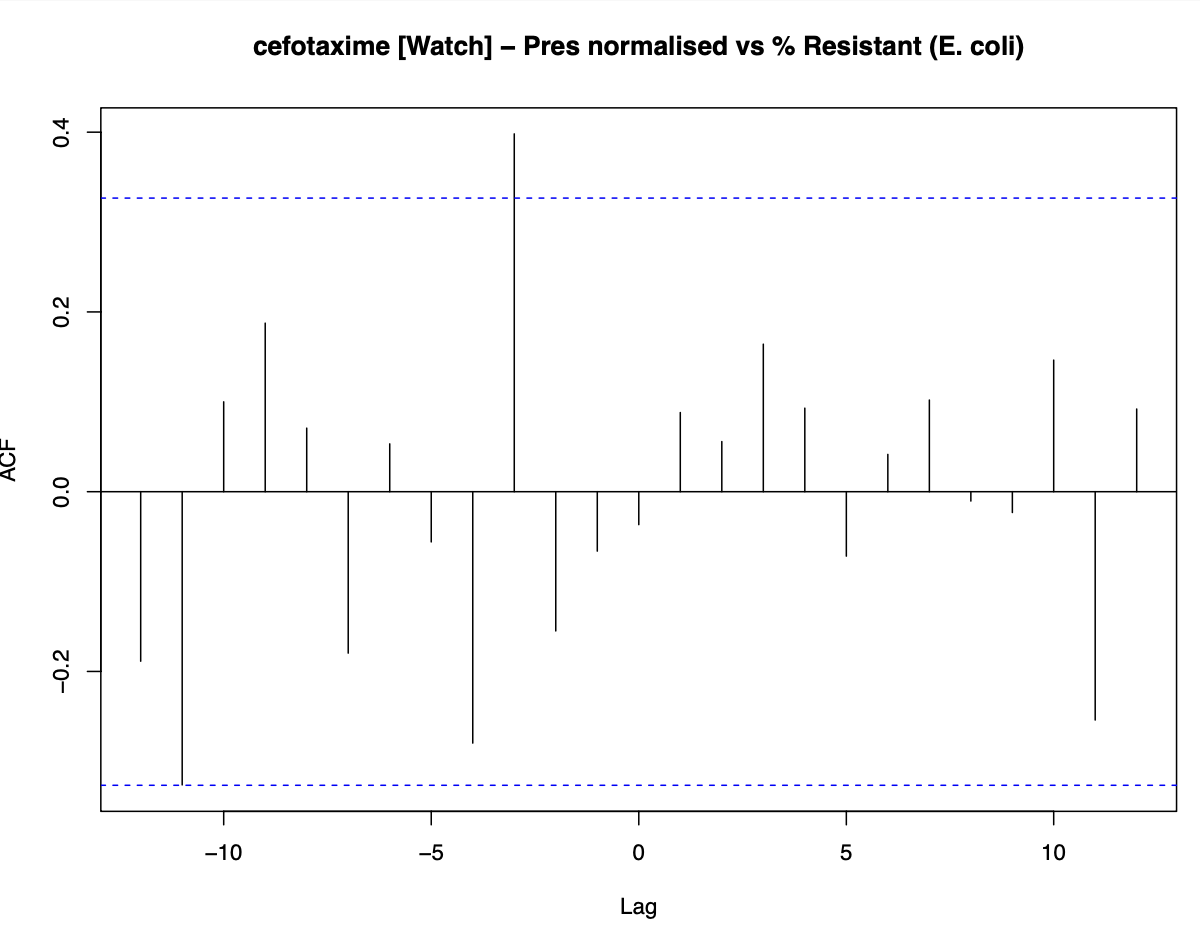

**Supplementary Figure 8: Pre-whitened cross-correlation between normalised cefotaxime prescriptions and urinary *E. coli* cefotaxime resistance.** For panels labelled A vs B, negative-lag peaks indicate that A preceded B. The plot shows the cross-correlation function between monthly normalised prescriptions and the monthly percentage of cefotaxime-resistant urinary E. coli isolates.
